# Dynamic Graph Representation Learning for Data-Driven Huntington’s Disease Staging: Evaluation Against Existing Embedding Methods and State-Space Models

**DOI:** 10.64898/2026.06.27.26355575

**Authors:** Lubna Mahmoud Abu Zohair, Hind Zantout, Alan Gow, John R. Woodward, Michael A. Lones, Marta Vallejo

**Author notes:** Corresponding author. Email address (Lubna Mahmoud Abu Zohair).

## Abstract

Huntington’s disease (HD) presents a heterogeneous neurodegenerative course, with motor, cognitive, and functional symptoms progressing differently across individuals. This atypical progression complicates the definition of discrete disease stages, hindering understanding of disease trajectories, timely patient care, and therapy development. Consequently, current clinical staging systems rely heavily on clinician-defined, domain-specific criteria and fixed clinical measurement boundaries for stage assignment, reducing objectivity and often leading to overlapping clinical measurements across stages. While machine learning methods can help, existing approaches cannot fully capture complex temporal relationships within and across patients. We propose URL-STFN, a dynamic graph-based representation learning model that encodes both inter- and intra-patient temporal patterns from longitudinal clinical measures. We then evaluate disease stages formed through clustering and stability analysis of URL-STFN latent representations, and compare them with representations obtained from conventional embedding approaches. We further benchmark these clustering-based stages against states derived from conventional temporal models, including DHMM. We hypothesize that clustering URL-STFN latent representations enables identification of HD stages with reduced overlap in clinical measurements. The proposed framework is evaluated using 1,477 clinical visits from the Enroll-HD dataset, a large longitudinal cohort with repeated clinical assessments. For staging, we used 44 clinical measurements spanning motor, cognitive, and functional domains. URL-STFN identifies clinically meaningful HD stages consistent with established disease progression while reducing overlap in clinical feature values compared with DHMM-derived and clinical staging approaches. These findings highlight the potential of a dynamic graph-based representation learning and clustering framework to support more objective, data-driven, and precise HD staging.

**Highlights:**

- A novel dynamic graph captures inter- and intra-patient HD progression latent.
- Clustering captured patterns identified clinically meaningful HD disease stages.
- Discovered stages showed reduced clinical feature overlap across clusters.
- Dynamic graph-based staging outperformed conventional embeddings and DHMM staging.
- Clustering multidomain latent features revealed new clusters beyond Shoulson–Fahn.

## 1. Introduction

Understanding disease progression is particularly challenging in neurodegenerative diseases (NDDs), like Huntington’s disease (HD), which exhibit abnormal dynamic changes in clinical symptoms and biomarkers [1, 2]. HD is a monogenic, autosomal dominant disorder caused by an expansion of cyto-sine–adenine–guanine (CAG) trinucleotide repeats in the HTT gene, leading to progressive neurodegeneration that affects motor, cognitive, and functional brain domains [3]. With variability in genetic expression among individuals, there is a corresponding variability in age at onset, symptom trajectories, and decline rates [3]. This multi-domain and longitudinal heterogeneity complicates disease course understanding, monitoring, and therapeutic stratification. Several clinical frameworks have been developed to characterize HD progression, reflecting the complexity of its motor, functional, and biological manifestations. Traditionally, clinical diagnosis of manifest HD relies on the Diagnostic Confidence Level (DCL) of the Unified HD Rating Scale (UHDRS), where manifest HD is defined as DCL=4, corresponding to 99% diagnostic confidence based on unequivocal motor abnormalities assessed by a clinical expert [4]. Functional staging complements DCL-based diagnosis by evaluating daily living and occupational capacities using the Total Functional Capacity (TFC) scale of the UHDRS. The Shoulson–Fahn (or TFC-based) staging system classifies individuals into five stages based on functional capacity [5]. Recognizing the need for a more comprehensive multidomain approach, the HD Integrated Staging System (HD-ISS) was recently introduced [6, 7]. This system integrates genetic expansion, imaging biomarkers (putamen and caudate volumes from MRI), UHDRS clinical signs (total motor score (TMS) or the symbol digit modalities test(SDMT)), and functional decline (TFC and Independence Scale scores) into a biologically informed framework that reflects disease progression across the lifespan. HD-ISS stage 0 includes individuals with ≥ 40 CAG repeats but no additional disease evidence. Stage 1 requires biological evidence such as caudate or putamen atrophy. Stage 2 combines these biological markers with observable clinical symptoms, and Stage 3 additionally includes functional decline.

Despite advances in HD staging, existing approaches are still limited. Most are domain-specific, typically relying on motor or functional measures such as DCL and TFC, and need clinician’s qualitative assessment of patient status (particularly in the commonly used DCL staging system) [8, 7, 9]. Or, they rely on clinician consensus-derived feature cutoffs to define diagnostic and disease stage thresholds. For example, transitions in the HD-ISS are operationalized using specific quantitative thresholds in imaging and clinical assessments to distinguish Stage 1 (biomarker evidence) from Stage 2 (clinical signs), and Stage 2 from Stage 3 (functional decline) [6, 7]. Similarly, the TFC-based staging system classifies individuals with HD into stages using predefined TFC score ranges [5]. Such threshold-based categorizations may introduce boundary effects and fail to resolve intra-stage heterogeneity [8]. Moreover, clinical assessments used for staging are susceptible to interrater variability, which has been documented in HD motor and functional evaluations [10, 11]. These limitations motivate complementary approaches that discover progression patterns directly from objective longitudinal clinical measurements or biomarkers, enabling a more precise and unbiased understanding of disease progression without reliance on fixed categorical rules [12].

Despite the availability of large longitudinal datasets such as Enroll-HD [13], current computational tools often struggle with the irregular, high-dimensional, and temporally evolving nature of clinical records. Classical machine learning approaches have commonly been applied to classify existing stages or progression patterns, and typically either: (1) work directly on static features but struggle to capture latent temporal dependencies and/or intra- or inter-feature relationships [12, 1, 14, 15, 16] or (2) rely on hand-crafted features based on pre-defined clinical or mathematical assumptions about variable relationships [17, 12].

Conventional deep learning models, such as convolutional neural networks (CNNs) and recurrent neural networks, capture complex patterns more effectively than traditional feature engineering. However, they operate on fixed input formats (sequences or grids) and do not inherently model evolving relational structures. In contrast, graph-based approaches explicitly encode relationships as part of the data structure, enabling the modeling of non-Euclidean and dynamically evolving dependencies [18, 19]. We hypothesize that this explicit structure can better capture both intra- and inter-patient dynamics over time, complementing implicit representations and improving the modeling of complex disease trajectories.

Existing graph applications in healthcare have largely focused on static graphs or short-term prediction tasks. For instance, Subgraph Neural Networks (SubGNNs) model disease phenotypes as static subgraphs derived from ontologies for classification tasks [20]. Dynamic graph approaches such as Multi-scale Spatiotemporal Dynamic GNNs (MSTD-GNNs) and continuous-time dynamic graphs (CTDGs) incorporate temporal information to enhance predictive performance for outcomes such as mortality or diagnosis [21, 22]. While these models integrate temporal signals, they remain primarily prediction-oriented and emphasize immediate or historical dependencies rather than long-term latent progression structure.

To date, no prior work has fully applied dynamic graph-based representation learning to model longitudinal clinical features in NDDs for unsupervised tasks, particularly for disease stage discovery, despite recent advances in graph explainability methods [23, 24, 25]. Moreover, it remains an open question whether node representations can be more meaningful for disease stages identification by jointly learning (i) generalized temporal patterns that capture dependencies across all time snapshots and (ii) time-specific node structures at each observation point. We hypothesize that such a unified modelling of global and local temporal structures can yield richer representations than static graph approaches or existing dynamic methods that primarily propagate information from past states. The applicability of the proposed representation learning approach to staging Parkinson’s progression using CSF biomarkers has been previously demonstrated [26]. Initial results from its application to HD are reported in [27, 28]; in this paper, we present a more in-depth study that compares against alternative representation learning methods, including competing deep learning approaches, and a direct comparison with disease stages derived from state-space modelling frameworks, which represent the state-of-the-art in this domain.

## 2. Methodology

Patient records corresponding to age ranges common to all participants were included. Patients’ longitudinal records were then represented as an age-based dynamic graph (denoted by G_Age1-G_AgeT in Figure 1), where each graph snapshot corresponds to a specific age, and nodes represent individual patients, with features representing their clinical measurements at that age. A temporal sequence of such graphs was constructed, capturing the evolution of patient profiles across consecutive ages. Age was chosen as the grouping anchor, not to claim the age as a key reason of abnormal evolvement but because it is one of the key risk factors to disease progression in HD and other neurodegenerative disorders, with progression often analyzed relative to chronological age and age at symptom onset [29, 30, 17]. This formulation enables the discovery of distinct disease stages that patients may variably traverse despite belonging to similar age ranges.

**Figure 1:**
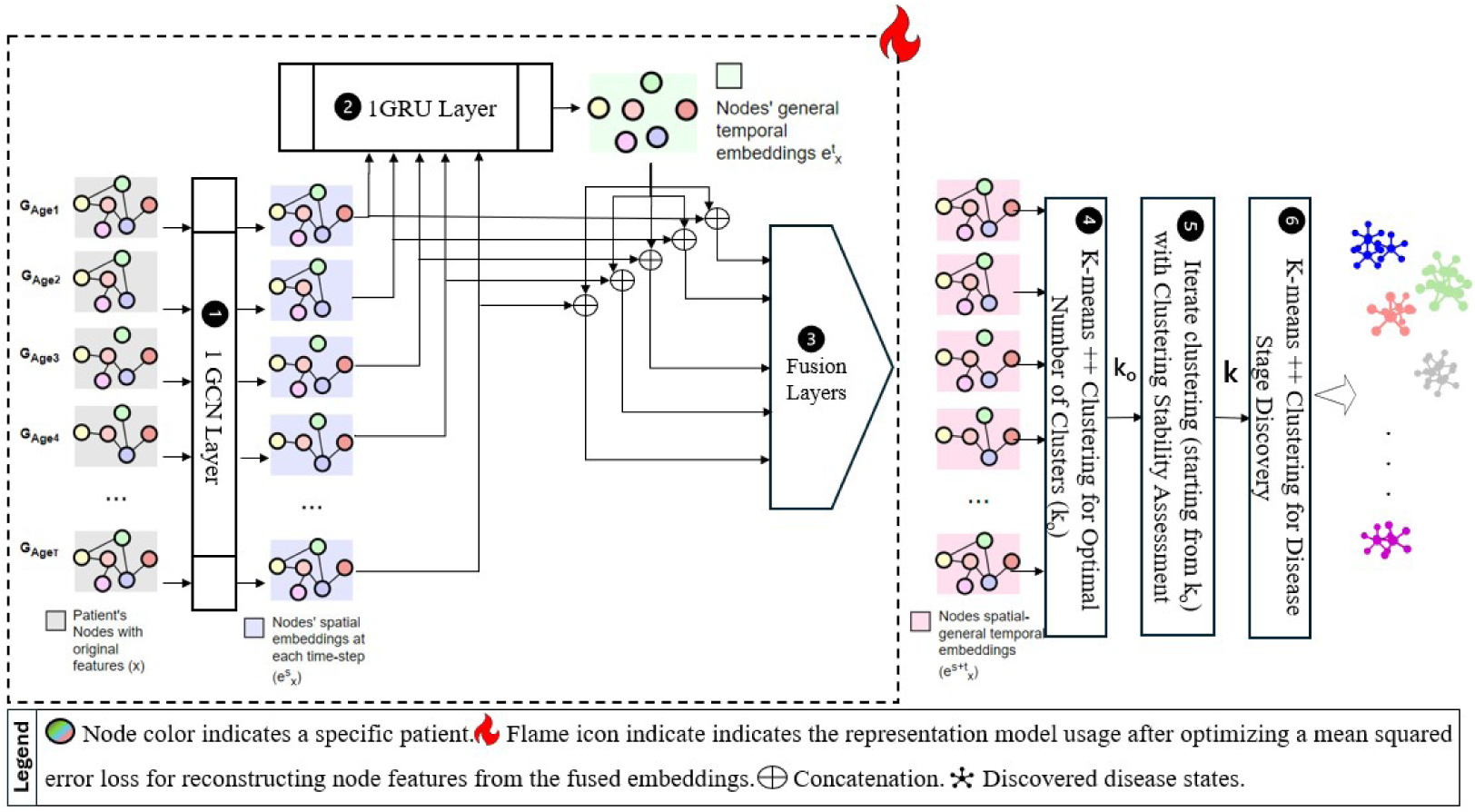
Overview of the Proposed Dynamic Graph Model Architecture. *G_Age_*_1−_*_AgeT_* represent the graphs constructed for each unique age in the longitudinal clinical visits’ dataset. Each node colour represents a unique patient who attended a clinical visit at the corresponding graph age. Missing nodes indicate time points where patients did not attend a clinical visit. The letter *x* represents the original features, *e* for the features’ embeddings. Superscript *S* refers to the spatial embeddings and superscript *t* denotes the temporal embeddings generated for each patient

Edges within each graph were defined using cosine similarity between patients’ clinical feature vectors, capturing intra-age similarity in clinical profiles. Cosine similarity was selected for its computational efficiency in high-dimensional spaces, robustness to variability in feature value ranges, robustness to sparse data, and effectiveness in comparing embedding vectors [31]. In addition, preliminary empirical assessments indicated that feature representations and clustering performance were comparable to those obtained using the Euclidean distance metric. Edges were added to connect patients with highly similar features, retaining only meaningful relationships and reducing noise. Specifically, an edge was formed when the cosine similarity between two nodes exceeded a threshold; the average 90th percentile of pairwise cosine similarity scores across all age-based graph snapshots.

### 2.1. URL-STFN Model Architecture

The dynamic evolution of patients’ clinical features (*x*) are represented as a series of age-based graph snapshots, denoted by *G_Age_*_1−_*_AgeT_* in Figure 1. Inspired primarily by researcher work in [32], an undirected graph was constructed at each age (*t*) in the longitudinal dataset (based on the age feature), as illustrated in Figure 1. This graph is annotated by

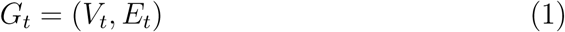

where:

- *V_t_*denotes the set of patient nodes at age *t*. Each node has features *x_i,t_* ∈ R*^d^*, representing the clinical measurements of patient *i* at age *t*. The feature matrix is defined as **X***_t_* ∈ R*^n^*^×^*^d^*, where *n* is the number of patients (nodes) and *d* is the number of clinical features associated with each node.
- *E_t_*denotes the set of edges between nodes. *E_t_*captures similarity-based relationships between patient features within the same age graph. The graph structure is represented by an adjacency matrix **A** ∈ R*^n^*^×^*^n^*, defined as:

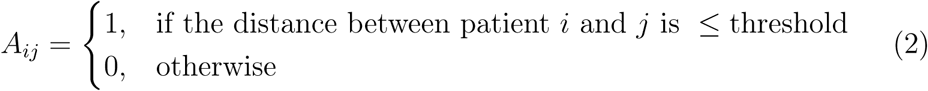

The structured dynamic graph was fed into the dynamic graph representation model (depicted in the structure surrounded be dotted rectangle in Figure 1), comprised of GCN, gated recurrent unit (GRU), and fusion layers. At each age-based graph, the GCN model learns spatial embeddings 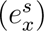 that encode the local relational context of each patient’s clinical features and aggregate information across direct neighbouring patients at age *t* (step 1 in Figure 1) [33]. Node embeddings are generated using a single GCN layer implemented via the GCNConv function in PyTorch Geometric. Unlike standard static GCN architectures, which typically employ multiple layers [33, 34], we intentionally adopt a single-layer design to emphasize direct neighborhood propagation. This choice mitigates over-smoothing effects associated with deeper graph convolutions, where repeated aggregation across higher-order neighborhoods can lead to homogenized node representations and reduced discriminative capacity across patients [35]. These embeddings are defined by Equation 3.

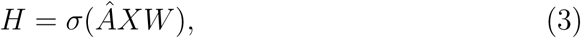

where: *H* represents the node embeddings, *̂A* is the normalized adjacency matrix, *W* is the learnable weight matrix, and *σ* is the activation function (ReLU) that combines the graph structure and node features *X* to compute the updated node embeddings.

These embeddings were subsequently organized as temporal sequences for each patient and passed through a recurrent layer to model longitudinal dependencies across visits (step 2 in Figure 1), enabling the model to capture historical and evolving clinical trajectory patterns 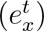. The spatial embeddings over time form this sequence *H_n_*_0_*, H_n_*_1_*, . . ., H_nt_*, and when fed into a GRU, a sequence of hidden states is produced; *Z_n_*_0_*, Z_n_*_1_*, . . ., Z_nt_*- where *Z_nt_* is the GRU’s hidden state for node n at time t. This is defined in Equation 4;

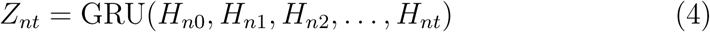

It is worth noting that the GCN and GRU components are modular; they can be substituted and tested with alternative graph or sequence models (such as Graph Attention Networks or Long Short-Term Memory models, respectively) without altering the overall architecture.

The spatial 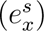 and temporal representations 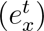 are then fused to produce a unified latent embedding 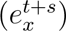, as depicted in step 3 in Figure 1. For each patient (n) at each clinical visit (t), the fusion process is summarized by Equation 5.

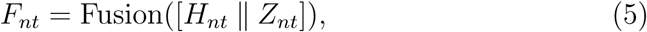

where: ‖denotes concatenation, and *F_nt_*∈ R*^N^*^×^*^d^* is the proposed model embedding for all nodes at every age *t*. Fusion corresponds to the fusion layers, which apply a fully connected layer to the concatenated embeddings *H_nt_* and *Z_nt_*, projecting them into a fixed fusion dimension (fusion_dim). A ReLU activation is applied to the output to introduce non-linearity and help capture complex relationship patterns. In the literature, fusion layers have been frequently employed to combine embeddings of features from multiple models or data modalities [36, 37]. The selection of these layers was made progressively, while assessing model training loss, resulting in the final configuration chosen based on achieving the lowest model loss.

The model components described above (highlighted by dotted rectangles in Figure 1) were jointly trained to reconstruct node feature representations rather than graph connectivity. Specifically, the model was optimized using a mean squared error (MSE) loss between the reconstructed and observed node features, in contrast to approaches that prioritize adjacency matrix reconstruction, such as [38]. This design choice was motivated by the study objective of learning embeddings that preserve clinically meaningful feature patterns. In patient graphs, where edges are often derived or uncertain, adjacency reconstruction can bias representations toward potentially noisy structural similarity [39]. Feature reconstruction instead emphasizes node’s features (patient-level clinical attributes)[39], which tend to provide more stable and clustering-relevant signals for phenotype discovery in attributed graph settings.

A transductive representation learning framework [40] was adopted, whereby embeddings for all nodes were learned during training and then directly utilized for downstream disease stage discovery. Following training, node embeddings were generated for each patient at each age-specific graph, as illustrated by the highlighted graphs in Figure 1. These embeddings were subsequently used as inputs to the downstream clustering algorithm. The dimensionality of the latent space was determined via grid search, with the optimal configuration selected based on the lowest training loss and convergence stability.

### 2.2. Conventional Features Representation Methods

To assess the effectiveness of the proposed URL-STFN model, we benchmarked it against competitive baseline implementations of conventional feature representation approaches identified in preliminary analyses. These included:

- Raw clinical measurements, without preprocessing.
- Kernel Principal Component Analysis (K-PCA). Non-linear dimensionality reduction for learning latent feature representations; selected after preliminary normality assessment using the Shapiro–Wilk test and Q–Q plots [41].
- Dense Autoencoder (Unsupervised) [42]. It is implemented as a shallow fully connected autoencoder with a single-layer encoder and a single-layer decoder, with mean squared error (MSE) for feature reconstruction.
- Graph Autoencoder (GAE) [38]. A single-layer static GCN model was implemented to restrict aggregation to first-order neighbors, to again mitigate over-smoothing effects that may arise from multi-hop message passing and lead to increasingly similar node representations [43]. The reconstruction loss of the adjacency matrix is computed using a cross-entropy function. This static graph model does not capture temporal patterns while learning the node representation.
- Temporal GCN (T-GCN) [44]. This is similar to the URL-STFN component implemented to capture the general temporal pattern. However, the node embeddings at each graph snapshot were extracted from GRU hidden states (which accounts for both local and historical temporal patterns only).
- Graph Convolution Embedded Long Short Term Memory (GC-LSTM)[45]. The model predicts the adjacency matrix at every age graph by minimizing the L2 (Frobenius) distance between the predicted probability matrix and the true adjacency matrix. Similar to T-GCN, the embeddings account for both local structural and current and historical temporal patterns only (unlike URL-STFN that consider historical and future temporal patterns while representing node features at every age).

In addition, to assess whether edge structural information contributes to the proposed model’s performance, we replaced the GCN component in the proposed architecture with a MultiLayer Perceptron (MLP) and evaluated the resulting model variant. This comparison informs the necessity of the structural features of the graph for the representations of the clinical visit learned at each time step (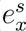 in Figure 1).

### 2.3. Comparative Evaluation Strategy

To evaluate the representation learning methods a hybrid clustering evaluation strategy was adopted because the true number and composition of the target clusters are unknown. After extracting the representations from the tested models, they were fed to the *KMeans* + + algorithm, known to provide statistical assurance on the quality of clustering [46]. First, the internal validity metrics that quantify intrinsic cluster structure (such as cohesion and separation) were used. This includes Silhouette score (SS), Davies–Bouldin index (DB), and Calinski–Harabasz index (CH) [47, 48, 49]. HD disease stages are not known and prior studies have demonstrated that internal indices alone cannot universally capture the true cluster structure across datasets [50]. However, when approximate external information is available (or clinical proxy variables like TFC- or DCL-based staging system), external agreement measures such as Adjusted Rand Index (ARI) can be used as validation signals. While these proxies do not constitute true ground truth, they provide a means to assess whether learned representations align with known clinical organization. This hybrid approach follows established recommendations in the clustering validation literature [50]. All models were evaluated across ten randomly initialized seeds to ensure the robustness and consistency of performance. In addition, a grid search was conducted to identify the optimal latent dimensionality from the set 4, 8, 16, 32, 64 for all evaluated models. This process included tuning the embedding dimensions of URL-STFN (i.e., 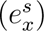,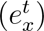, and 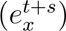), with the final configuration selected based on convergence to the minimum loss value. Preliminary tests conducted with higher dimensions resulted in degraded training loss and clustering performance. This analysis was conducted across three data subset sizes, including cohorts of patients with longitudinal clinical visits conducted at common ages, comprising 5, 6, and 7 visits per patient. The optimal representation model was selected based on achieving consistent and strong internal clustering validity while also demonstrating agreement with existing clinical staging proxies, both across seeds and data subsets. The best model was subsequently used to identify HD stages.

### 2.4. Disease Stage Discovery Framework

Building on the selected optimal representation, the learned embeddings were propagated through subsequent analysis steps (steps 4–6 in Figure 1) to identify latent disease stages and evaluate their clinical relevance. Conventional clustering performance metrics primarily optimize separability between groups, potentially overlooking intermediate or transitional patterns that may carry meaningful structure. In unsupervised settings without ground-truth labels, this limitation is further compounded by uncertainty in the underlying data structure and the possibility of missing patterns due to limited sample size [51]. We therefore hypothesize that clustering stability assessment can complement conventional clustering evaluation metrics by identifying statistically distinct subgroup structures beyond the most parsimonious solution, beyond the number of clusters with the highest internal validity score.

The final fused latent representations produced by the fusion layer were clustered using *K*-means++ (step 4 in Figure. 1), with grid searching over different numbers of clusters (*k*) to identify the *k_o_* that maximizes the silhouette score (SS) (step 4 in Fig. 1). The best *k_o_*was then used as the initial clustering solution, after which, cluster stability analysis was used to investigate whether additional stable and meaningful clusters exist. *k_o_* was incrementally increased, and for each value of *k_o_*cluster stability was assessed by comparing cluster assignments derived from 10 bootstrap samples (each containing 80% of the dataset, sampled with replacement) to those obtained from clustering the full dataset (step 5 in Figure 1). This comparative assessment was performed using two criteria: Optimal Transport Alignment (OTA; based on the Hungarian algorithm) [52], which is commonly applied in biomedical and omics data for subgroup discovery, and the Bootstrap Jaccard coefficient (BJ) [53]. Mean stability values above a threshold of 0.8, as suggested by [51], were considered indicative of robust subgroup structures. Once stability fell below this threshold, the exploration was terminated, and the last stable solution (*k*) was selected as the final number of clusters. This indicates that, while *k_o_* clusters provide the most parsimonious separation, models with up to *k* clusters still form meaningful and robust subgroup structures that are unlikely to arise from random data variation.

The resulting clusters were statistically validated using permutational multivariate analysis of variance (PERMANOVA) to assess multivariate separation [54], while differences in individual clinical measurements across the discovered clusters were examined using the Kruskal–Wallis test [55], with Bonferroni correction applied to adjust for recurrent comparisons across features, with statistical significance set at p-value*<*0.05. Finally, the identified disease stages were characterized through visual analysis of clinical profiles and temporal progression patterns, contextualized with findings from the literature. Feature distributions and their overlap across stages were evaluated against established staging systems. Visualizations, including boxplots, radar charts, bar and line graphs, and heatmaps, were used to illustrate (i) motor, functional, and cognitive clinical profiles, (ii) temporal patterns of cluster transitions, and (iii) the degree of overlap in clinical measurement values between the derived clusters, compared with that observed in existing staging frameworks.

### 2.5. Huntington Stages by State-Space Model

We evaluated whether the proposed unsupervised ML pipeline provides an improved profile of the characteristics of disease states compared to state space models, which have been widely used in the literature to characterize the progression of NDDs [56, 57]. The selected clinical measurements are ordinal and do not follow the normal distribution pattern. Therefore, we employed a probabilistic state-space modelling framework based on Discrete Hidden Markov Models (DHMM), a well-established class of generative models used to uncover complex states in evolving latent processes [58, 59]. The model assumes that there are hidden states (underlying disease stages) that change over time, and the next state depends only on the current one. At each age, clinical measurements were observed, and their values were assumed to be mainly explained by the underlying hidden disease stage rather than by direct relationships between the measurements themselves.

Let 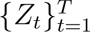 denote the unobserved disease states and 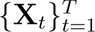 the observed feature vectors, **X***_t_* = (*X_t_*_1_*, . . ., X_t_*_44_). The hidden states’ dynamics follow a first-order Markov chain:

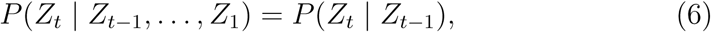

with transition probabilities

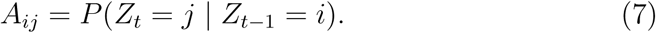

Conditional on *Z_t_*, features are independent and ordinal:

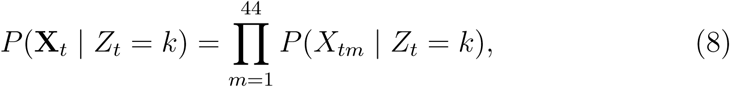

Model parameters were estimated by maximum likelihood using the Ex-pectation–Maximization (EM) algorithm, while the number of hidden states *k* was selected based on the Bayesian Information Criterion (BIC) evaluated across five random initializations to ensure stability. This approach follows prior work in disease progression modelling, where EM facilitates efficient likelihood maximization and convergence, and BIC provides a robust criterion to prevent overfitting [56, 57].

We evaluated the staging derived by the DHMM by examining the characteristics of clinical feature values and their overlap between the derived clusters. Based on these results, the staging method demonstrating the most robust and consistent patterns was recommended.

### 2.6. Dataset and Preparations

Data used in this work were generously provided by the participants in the Enroll-HD study and made available by the CHDI Foundation, Inc. Enroll-HD is a global clinical research platform designed to facilitate clinical research in HD. Core datasets are collected annually from all research participants as part of multi-center longitudinal observational study. Data are monitored for quality and accuracy using a risk-based monitoring approach. All sites are required to obtain and maintain local ethical approval. In particular, patients’ longitudinal records in the Enroll table in Enroll-HD PDS7-R1 clinical dataset were mainly utilized in providing the concept of the proposed model. This table initially comprised around 30,511 unique participants and 129,537 recorded visits [60, 61, 13].

We limited the analysis to patients who had records across at least four consecutive annual clinical visits. This selection helps ensure that potential disease states or transitions are not missed due to gaps in yearly visits. By requiring shared longitudinal coverage across patients, all individuals are evaluated over comparable time points, reducing the bias caused by missing or irregular observations. For example, if Patient 1 has clinical visits recorded at ages 42, 43, 44, and 45, then Patient 2, Patient 3, . . ., Patient n should also have visit records at these same ages (42–45). Only patients with observations at these common sequential ages are retained for the analysis. The dataset preparation steps and preprocessing procedures, summarized in Fig. 3, involved the following steps:

- Participants classified as family controls or genotype-negative were excluded. Filtering was performed using the common identified ’subjid’ across the “enroll.csv” and “participant.csv” tables. Individuals were selected based on the ’hdcat_0’, retaining only those with premanifest or manifest motor stages.
- A total of 44 motor, cognitive, and functional clinical measurements were selected. These were chosen because they are commonly utilized in prior clinical staging studies [62, 5, 17], enabling better interpretation of the discovered disease stages. Aggregated features, such as CAP score and composite variables derived from clinical assessments (specifically TMS: motscore, TFC: tfcscore, and DCL: diagconf), were excluded from direct input to the models used for representation learning or clustering. These features were instead considered when discussing the characteristics of the disease stages derived from the selected granular clinical measurements.
- Handling Missing Data. Temporal records that have all features missing (NaN) were dropped. For features with less than 40% missing values, missing data were explicitly imputed with a constant value (−1) [63], while those exceeding this threshold were excluded. While no universal threshold exists for feature exclusion, prior work shows that high levels of missing data (exceeding 40%) can increase estimation uncertainty and introduce bias when simple imputation approaches are used, accordingly, features with very high missingness are typically considered suitable only for hypothesis-generating analysis [64]. Therefore, to ensure meaningful representation learning for HD staging, such features were excluded. All clinical variables used for clustering are non-negative ordinal measures; therefore, -1 lies outside the physiologically plausible range and serves as an explicit indicator of missingness rather than a valid measurement. This strategy preserves missingness information while avoiding imputation-induced bias, dilution, or instability that can arise from low-performing machine learning–based imputations.
- Feature Standardization. The Enroll-HD dataset is known to contain clinical features measured on heterogeneous scales and to exhibit skewed distributions and extreme values. To mitigate the variable scaling influence, the StandardScaler function from the scikit-learn package. was applied to standardize features to zero mean and unit variance, ensuring that variables measured on different scales contribute comparably to the analysis [11, 65].

**Figure 2:**
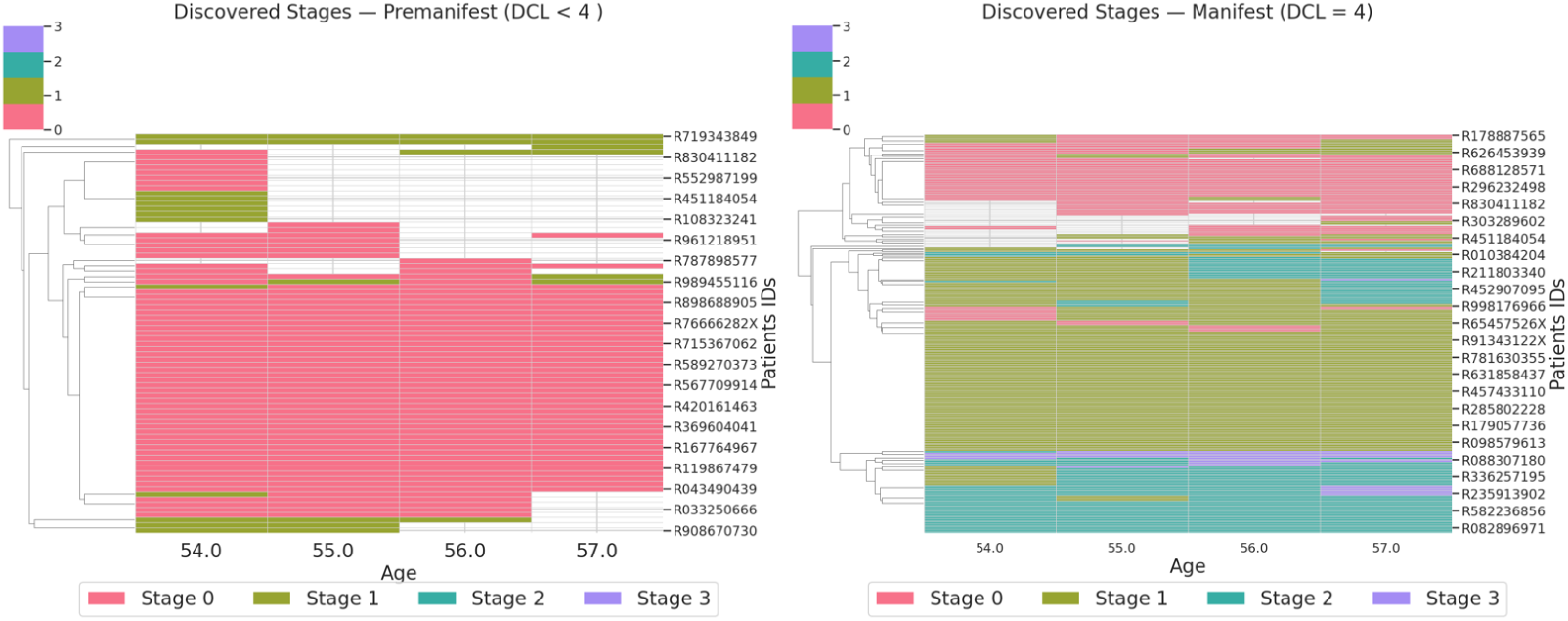
Heatmaps show the distribution of discovered clusters across patient visits. The clinical premanifest heatmap (DCL *<* 4) displays visits occurring in the premanifest stage, while the manifest panel (DCL = 4) shows visits classified as manifest, with colours indicating the corresponding cluster assignment. Rows represent subjects and are hierarchically clustered to highlight similarities in longitudinal trajectories; columns represent common ages at visit. Colours correspond to actual cluster numbers (legend), with white cells indicating patients’ visits that do not belong to the current stage depicted in the heatmap.

**Figure 3:**
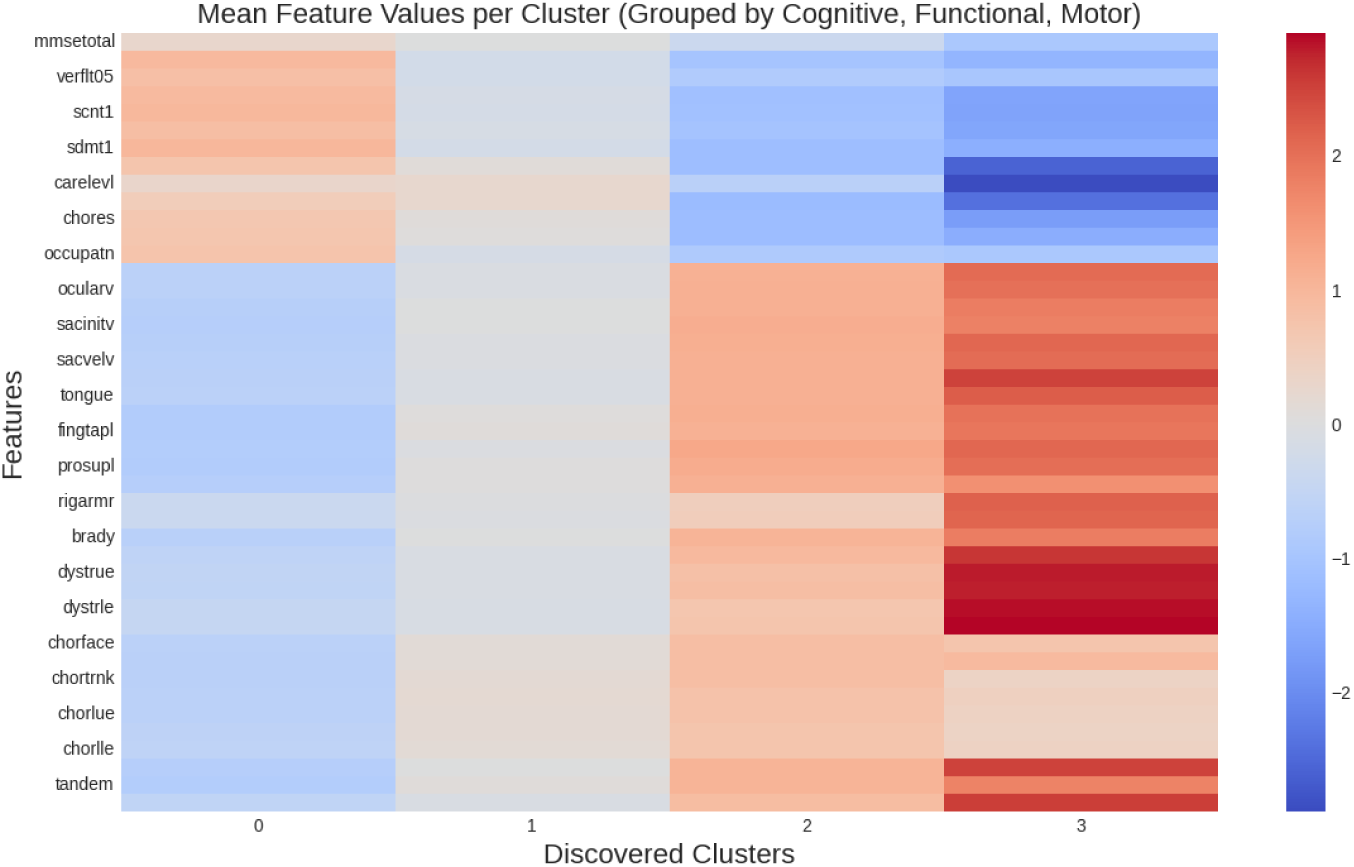
Heatmap shows the mean values of all features across the discovered clusters. Features in the Y-axis grouped by domain: cognitive (top), functional (middle), and motor (bottom). This visualization highlights domain-specific patterns and the variation of feature severity across clusters.

## 3. Results and Discussion

The selected subset of Enroll-HD comprised longitudinal records from 302 patients, with 1,477 clinical visits and 44 clinical features (described in Table A in the Supplementary_File_1). Most participants were in the manifest stage (80%), were in their 50s, and most commonly had a CAG repeat length of 42. As shown in Table 1 approximately half of the observations were used as a subset of the main dataset for comparative evaluation of the representation learning models. The full dataset, which includes at least four clinical visits per patient, was subsequently restricted to the four common clinical visits and used to infer underlying disease stages using the best-performing representation learning model.

**Table 1:** Clinical characteristics of the Enroll-HD datasets subsets used for uncovering HD disease stage (Main Dataset, with at least four common ages) and representation learning models’ comparison (with 5, 6, and 7 common longitudinal visits).

| Characteristic | Main Dataset | 5 Visits | 6 Visits | 7 Visits |
| --- | --- | --- | --- | --- |
| Number of participants | 302 | 154 | 84 | 31 |
| Number of observations | 1208 | 770 | 504 | 217 |
| Age (range) | 54.0–57.0 | 53.0–57.0 | 52.0–57.0 | 51.0–57.0 |
| CAG repeats (mean ± SD) | 42.5 ± 1.9 | 42.6 ± 1.9 | 42.7 ± 1.7 | 42.7 ± 1.9 |
| Premanifest (count, %) | 234 (19.4%) | 176 (22.9%) | 119 (23.6%) | 33 (15.2%) |
| Manifest (count, %) | 972 (80.5%) | 593 (77.0%) | 384 (76.2%) | 183 (84.3%) |

### 3.1. Comparative Evaluation of Representation Models

Across 10 random seeds and varying dataset sizes, Table 2 demonstrates that URL-STFN consistently achieves robust clustering performance while maintaining a compact latent dimensionality of 4 across all representations (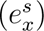,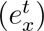, and 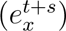; see Figure 1). In terms of primary internal clustering metrics, URL-STFN consistently yields high SS with low variance (0.665±0.026, 0.658±0.021, 0.678±0.029 for dataset sizes 7, 6, and 5, respectively), alongside stable DB scores (0.575±0.041, 0.578±0.038, 0.571±0.038). This indicates well-separated and compact clusters across all dataset sizes.

**Table 2:** Mean ± standard deviation (SD) of clustering performance metrics computed over 10 random seeds for representations learned by different algorithms, evaluated on patient dataset subsets with varying longitudinal depth (5, 6, and 7 longitudinal records). Underlined font indicates methods that consistently perform best/are consistent across all metrics for a given dataset size.

| Method | Best Dim. | Silhouette ↑ | DB ↓ | CH ↑ | ARI_DCL ↑ | ARI_TFC ↑ |
| --- | --- | --- | --- | --- | --- | --- |
| **Dataset Size = 7** |  |  |  |  |  |  |
| URL-STFN | (4,4,4) | <u>0.665 ± 0.026</u> | <u>0.575 ± 0.041</u> | 455.180 ± 82.802 | -0.108 ± 0.001 | 0.249 ± 0.013 |
| MLP | (4,4,4) | 0.658 ± 0.140 | 0.519 ± 0.226 | 575.845 ± 363.787 | 0.068 ± 0.073 | 0.207 ± 0.054 |
| GC-LSTM | (4,4) | 0.471 ± 0.017 | 0.901 ± 0.075 | 163.528 ± 10.889 | 0.005 ± 0.006 | -0.014 ± 0.016 |
| T-GCN | (4,4) | 0.529 ± 0.053 | 0.723 ± 0.098 | 265.556 ± 90.162 | -0.002 ± 0.006 | 0.003 ± 0.005 |
| GCN | D=4 | 0.329 ± 0.016 | 1.110 ± 0.086 | 100.439 ± 8.773 | 0.006 ± 0.009 | 0.048 ± 0.021 |
| Dense AE | D=4 | 0.436 ± 0.039 | 0.855 ± 0.052 | 154.379 ± 38.997 | 0.040 ± 0.032 | 0.105 ± 0.038 |
| K-PCA Features | 17 | 0.6132 ± 0.000 | 0.4456 ± 0.000 | <u>2223.263 ± 0.000</u> | 0.0295 ± 0.000 | 0.1262 ± 0.000 |
| Raw Features | 2 | 0.404 ± 0.004 | 1.044 ± 0.011 | 159.956 ± 0.151 | -0.099 ± 0.002 | 0.163 ± 0.007 |
| **Dataset Size = 6** |  |  |  |  |  |  |
| URL-STFN | (4,4,4) | 0.658 ± 0.021 | <u>0.578 ± 0.038</u> | 1019.520 ± 134.449 | -0.044 ± 0.131 | 0.216 ± 0.010 |
| MLP | (4,8,4) | 0.631 ± 0.048 | 0.561 ± 0.091 | 1337.329 ± 613.895 | 0.181 ± 0.105 | 0.318 ± 0.039 |
| GC-LSTM | (4,4) | 0.495 ± 0.024 | 0.829 ± 0.030 | 425.583 ± 48.424 | 0.019 ± 0.014 | 0.037 ± 0.018 |
| T-GCN | (4,4) | 0.532 ± 0.030 | 0.752 ± 0.090 | 626.401 ± 126.825 | -0.002 ± 0.004 | -0.001 ± 0.003 |
| GCN | D=4 | 0.332 ± 0.006 | 1.168 ± 0.020 | 264.63 ± 11.32 | 0.170 ± 0.052 | 0.052 ± 0.019 |
| Dense AE | D=4 | 0.472 ± 0.041 | 0.862 ± 0.095 | 468.127 ± 85.656 | 0.152 ± 0.070 | 0.255 ± 0.060 |
| K-PCA Features | 6 | 0.6268 ± 0.000 | 0.4658 ± 0.000 | <u>1993.6361 ± 0.000</u> | 0.1599 ± 0.000 | 0.2695 ± 0.000 |
| Raw Features | 2 | 0.3203 ± 0.000 | 1.2374 ± 0.000 | 308.988 ± 0.000 | 0.0659 ± 0.000 | 0.1774 ± 0.000 |
| **Dataset Size = 5** |  |  |  |  |  |  |
| URL-STFN | (4,4,4) | <u>0.678 ± 0.029</u> | <u>0.571 ± 0.038</u> | 1655.19 ± 240.72 | -0.090 ± 0.006 | 0.240 ± 0.011 |
| MLP | (4,16,4) | 0.670 ± 0.096 | 0.509 ± 0.156 | <u>5895.173 ± 10331.441</u> | 0.139 ± 0.071 | 0.347 ± 0.032 |
| GC-LSTM | (4,4) | 0.554 ± 0.021 | 0.842 ± 0.040 | 858.775 ± 116.92 | 0.009 ± 0.019 | 0.039 ± 0.027 |
| T-GCN | (4,4) | 0.526 ± 0.044 | 0.761 ± 0.119 | 1035.312 ± 229.479 | -0.000 ± 0.004 | -0.001 ± 0.001 |
| GCN | D=4 | 0.375 ± 0.017 | 1.087 ± 0.042 | 523.454 ± 61.290 | 0.049 ± 0.013 | 0.096 ± 0.017 |
| Dense AE | D=4 | 0.531 ± 0.059 | 0.844 ± 0.040 | 403.79 ± 90.47 | 0.148 ± 0.102 | 0.255 ± 0.051 |
| K-PCA Features | 2 | 0.6107 ± 0.000 | 0.5481 ± 0.000 | 1691.0731 ± 0.000 | 0.2365 ± 0.000 | 0.3142 ± 0.000 |
| Raw Features | 2 | 0.350 ± 0.0004 | 1.163 ± 0.0007 | 604.295 ± 0.000 | -0.001 ± 0.000 | 0.189 ± 0.000 |

In contrast, competing methods such as MLP occasionally achieve comparable or even higher secondary metrics (ARI_TFC_ = 0.318 and 0.347 for sizes 6 and 5), but exhibit low internal clustering validity performance and substantially higher variance (with SS std = 0.140 and 0.096, DB std = 0.226 and 0.156), reflecting instability across seeds. Also, they did not maintain a unified latent representation across dataset sizes. Similarly, graph-based baselines (GC-LSTM, T-GCN, and GCN) consistently produce lower silhouette scores (*p* ≤ 0.554) and higher DB scores (≥0.723), indicating weaker clustering structure. While K-PCA achieves strong Calinski–Harabasz scores, its performance is less consistent and does not maintain a unified latent representation across dataset sizes. The dense autoencoder provides moderate, relatively stable performance but remains inferior to URL-STFN in both separation quality and consistency.

These results indicate that strong internal clustering quality does not always align with higher ARI scores. Both URL-STFN and MLP variants show comparable performance, but following prior literature, internal validity metrics were prioritized, treating ARI proxy stages as a secondary assessment. URL-STFN, with its consistently low-dimensional latent space, provides a robust candidate approach for capturing clinically meaningful patterns that delineate cluster-derived stages of HD. However, MLP-based representation merits further exploration in the future.

### 3.2. Discovered Disease Stages

We tested the main dataset (i.e. 302 patients records with 4 clinical visits each) using the candidate representation learning algorithm, URL-STFN, and identified a two-cluster solution (SS = 0.67, DB = 0.584, CH = 2250.1738), and these clusters were statistically distinct according to PERMANOVA analysis (*p*≤0.05). The stability of these clusters was assessed, and the potential for identifying more granular subgroups was evaluated using BJ and OTA criteria. Stability and significance were maintained up to four clusters using 10 bootstrap iterations with 80% of the dataset; however, at this higher cluster number, inter-cluster separation and intra-cluster compactness slightly declined (SS = 0.51, DB = 0.66, CH = 1528.97), indicating a trade-off between cluster granularity and overall clustering quality. The discovered clusters were mapped to longitudinal visits of patients categorized as premanifest (DCL*<*4) and manifest (DCL=4) according to the criteria of UHDRs [4], and visualized in heat maps (see the heat maps on the left and right in Figure 2). This visualization demonstrates patterns across clusters that appear to correspond with progression from premanifest to manifest disease stages. The cluster labels (0–3) do not reflect an intrinsic ordering from the clustering algorithm, as they are assigned arbitrarily in an unsupervised setting. For interpretability, clusters were ordered post hoc based on clinical characteristics and transition patterns observed in the data. Cluster 0 was observed predominantly during the premanifest phase, whereas Cluster 1 appeared in both premanifest and manifest visits but occurred more frequently following manifest conversion. In contrast, clusters 2 and 3 were largely confined to manifest visits. Together, these patterns suggest alignment between the identified clusters and clinical disease staging.

The progressive severity patterns across the discovered clusters are illustrated by standardized clinical feature visualizations (Figures 3 and 4); corresponding standardized means and 95% CIs are reported in Table B (Sup-plementary_File_2) for URL-STFN–derived stages and established staging systems (DCL-, TFC-, and HDISS-based). These show that cluster 0 corresponds to a relatively preserved clinical state, while clusters 1–3 exhibit progressively worsening motor, cognitive, and functional impairment. All features differed significantly across clusters (Kruskal–Wallis H-test, *p* ≤ 0.05), indicating that the clustering captures clinically meaningful gradients of disease severity.

#### 3.2.1. Clinical Profiles of Discovered Stages

Examining the actual mean values of all features, in addition to Figure 4, Table 3 shows the progression of feature values, revealing a clear worsening is observed from Cluster 0 to Cluster 1, further supported by violin plots of individual feature distributions (Figures A–C, Supplementary_File_1), where this pattern becomes more apparent. Together, these representations illustrate a consistent progression, with cognitive, functional, and motor measures declining progressively from Cluster 0 to Cluster 3.

**Figure 4:**
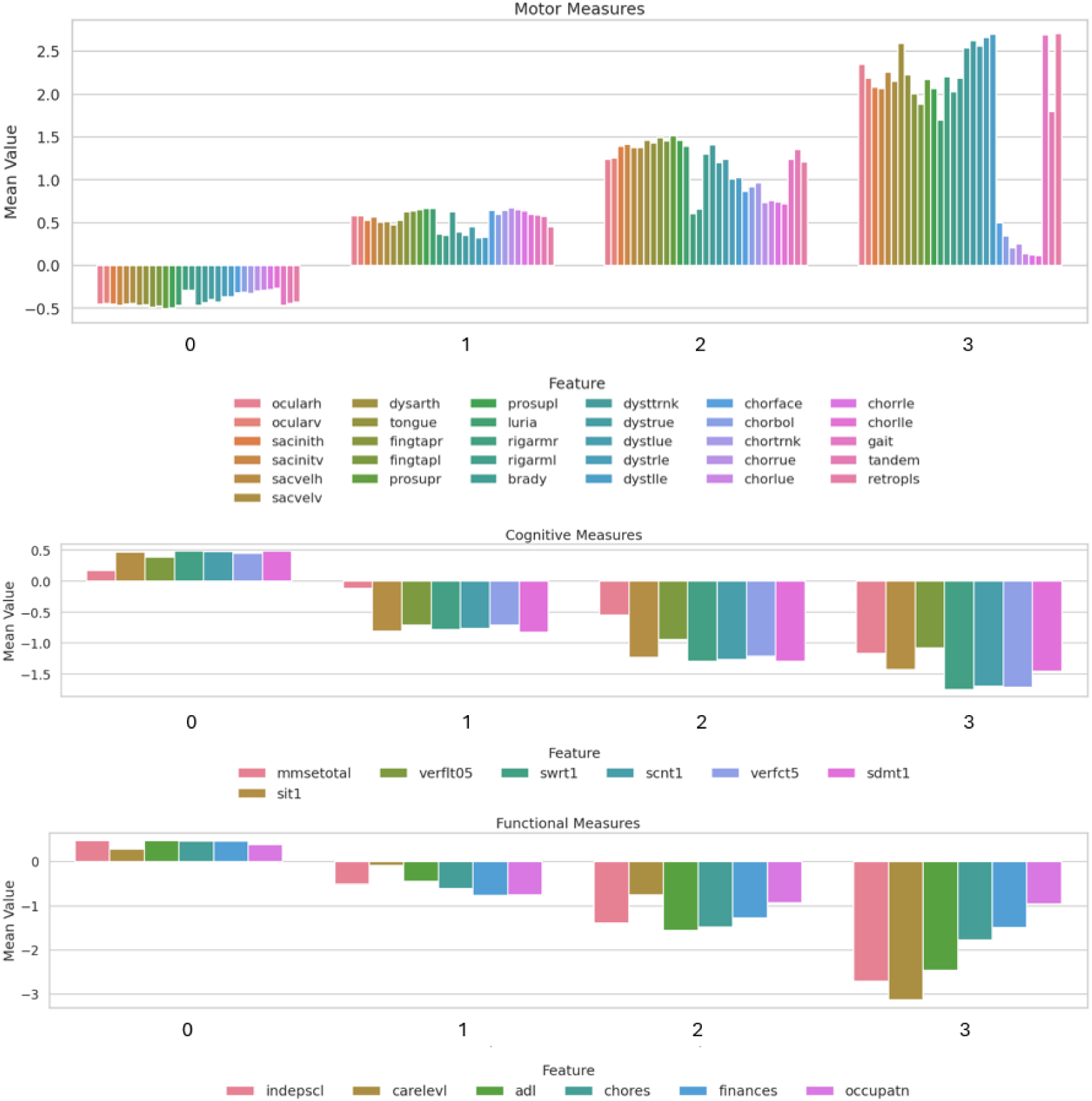
Bar graph for the standardized mean clinical feature values across the discovered clusters, grouped by motor (top bars), cognitive (middle bars), and functional (bottom bars) domains. Bars represent the mean z-scored values of each feature within each cluster, illustrating relative deviations from the cohort mean.

**Table 3:**
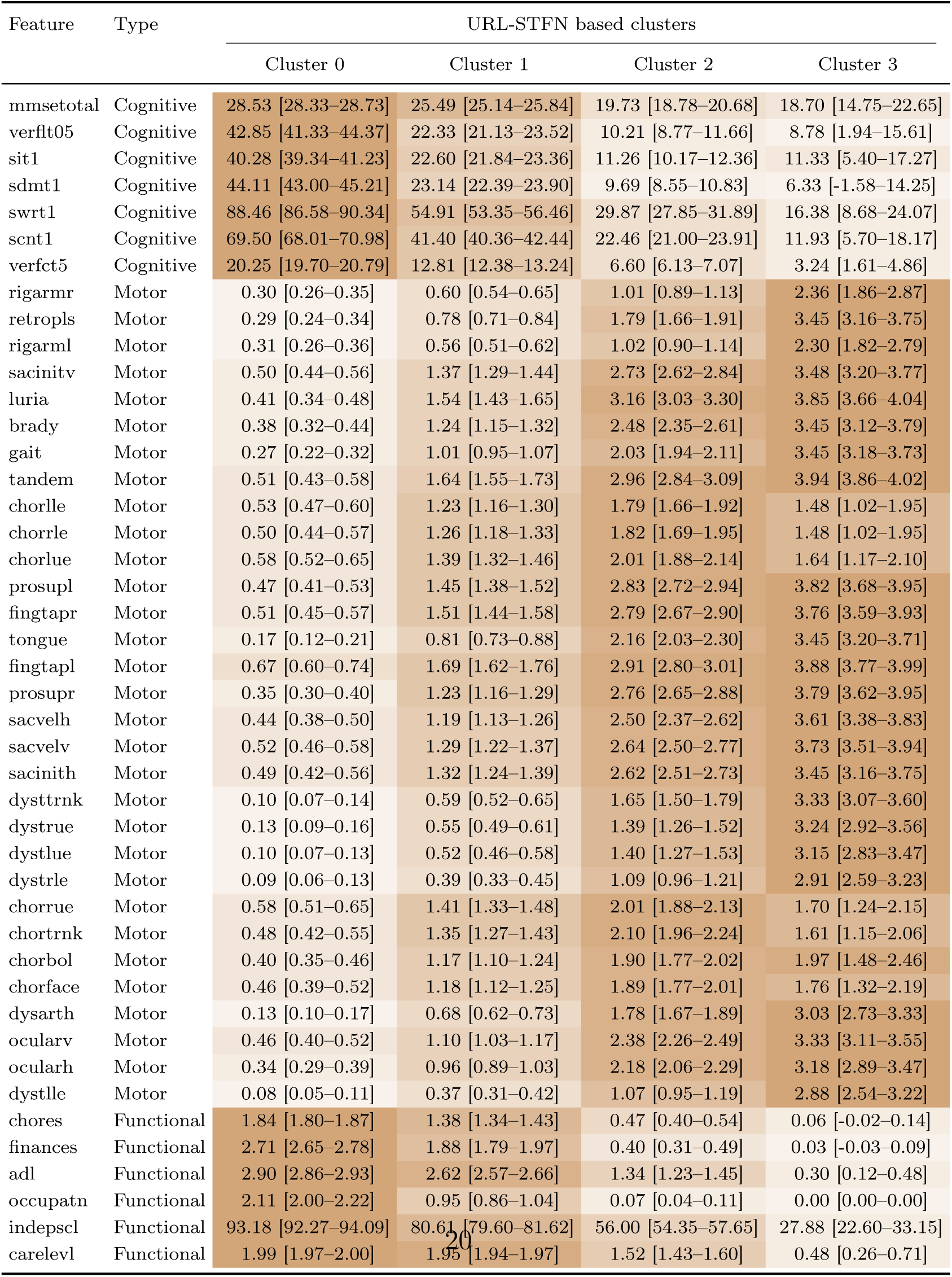
Mean (95% CI) of 44 features across URL-STFN clusters. Darker brown indicates higher values; for motor features, it reflects worse condition, while for cognitive and functional features, it reflects better condition, and vice versa.

For instance, looking at the cognitive features in Table 3, mini-mental state examination (MMSE) mean scores decreased from 28.53 in cluster 0 to 25.49 in cluster 1, 19.73 in cluster 2, and 18.70 in cluster 3. A similar decline was observed for the SDMT1, with mean [95% CI] scores of 44.11 [43.00–45.21], 23.14 [22.39–23.90], 9.69 [8.55–10.83], and 6.33 [-1.58–14.25] across clusters 0–3, respectively. Likewise, Verbal Fluency (letter fluency, verflt05) and Stroop Word Reading (swrt1) scores also progressively decreased across clusters. Additional cognitive measures are reported in Table 3 for completeness.

Building on the cognitive decline described above, functional capacity patterns across clusters (Figure 4 and Table 3) exhibited one of the steepest gradients. For example, Independence scale (indepscl) scores decreased from 93.18 [92.27–94.09] in Cluster 0 to 80.61 [79.60–81.62], 56.00 [54.35–57.65], and 27.88 [22.60–33.15] in Clusters 1–3, reflecting the progressive need for care. Similarly, activities of daily living (adl) scores declined from 2.90 [2.86–2.93] to 2.62 [2.57–2.66], 1.34 [1.23–1.45], and 0.30 [0.12–0.48] across clusters. These functional measures, together with the cognitive assessments, are highly relevant for prognosis, caregiver burden, and resource planning, as it likely marks the stage at which patients move from partial independence to sustained dependency.

In parallel, motor impairment increased steadily across clusters (see Figure 4): bradykinesia (brady) rose from 0.38 [0.32–0.44] in Cluster 0 to 1.24 [1.15–1.32], 2.48 [2.35–2.61], and 3.45 [3.12–3.79] in Clusters 1–3, while gait impairment increased from 0.27 [0.22–0.32] to 1.01 [0.95–1.07], 2.03 [1.94–2.11], and 3.45 [3.18–3.73], as recorded in Table 3, Postural instability and tandem gait impairment became particularly pronounced in later clusters, contributing substantially to functional decline. Similarly, oculo-motor abnormalities followed the same pattern, with vertical saccade velocity (sacvelv) increasing from 0.52 [0.46–0.58] in cluster 0 to 1.29 [1.22–1.37], 2.64 [2.50–2.77], and 3.73 [3.51–3.94] in cluster 3. There is another noteworthy chorea-related observation: hyperkinetic choreiform movements measurements (chorlle, chorrle, chorlue, chorrue, . . .) where severity increased from early to intermediate stages but appeared to attenuate in cluster 3 (see the top bar plot in Figure 4, and in numbers in Table 3).

### 3.3. Validation of URL-STFN Cluster-Based Staging Against Established HD Frameworks

Given that the calculated mean UHDRS TFC for Cluster 0 was 13.75 (Table B; Supplementary File 2), derived from the summed mean motor feature scores [66], this value is comparable to those reported in large longitudinal cohorts such as PREDICT-HD and TRACK-HD, where average TFC values at the point of confirmed motor diagnosis (DCL = 4) are typically around 15, albeit with substantial inter-individual variability (approximately 8–35) [67]. Notably, [66] classified such values within the manifest stage (DCL = 4).

For the discovered Clusters 1, 2, and 3, both the mean TMS and TFC scores (as shown in Table B) are consistent with the manifest phase as defined in established HD staging frameworks. Cluster 1 appears to align with a transitional phase between Shoulson–Fahn stages II and III, while Clusters 2 and 3 fall within the TFC-defined Stage III range [68]. This pattern suggests that the proposed clustering framework may further subdivide the conventional Stage III category into more granular sub-stages. Importantly, these sub-stages are characterized by distinct profiles across TFC and other clinical measures, indicating that the observed stratification is likely to reflect meaningful clinical heterogeneity rather than random variation.

The distinction between stages identified by the ML-based approach becomes more apparent when compared with established staging systems through the visualization of feature distributions across clusters. These distributions are summarized using mean standardized (z-score) values and corresponding variability in Figure 5, and median-based summaries in Figure 6. Looking at the main features, Figure 5 demonstrates how the features-value patterns of the discovered stages (outlined in red) are distinct but more closely aligned with TFC-based staging than with other established frameworks. Notably, the discovered stages maintain clear distinctions, with minimal overlap observed between stages across the examined features. Given that most clinical measurements were not normally distributed (Table C, Supplemen-tary_File_2), medians and interquartile ranges (IQRs) were used to characterize and assess stage-wise separation and overlap.

**Figure 5:**
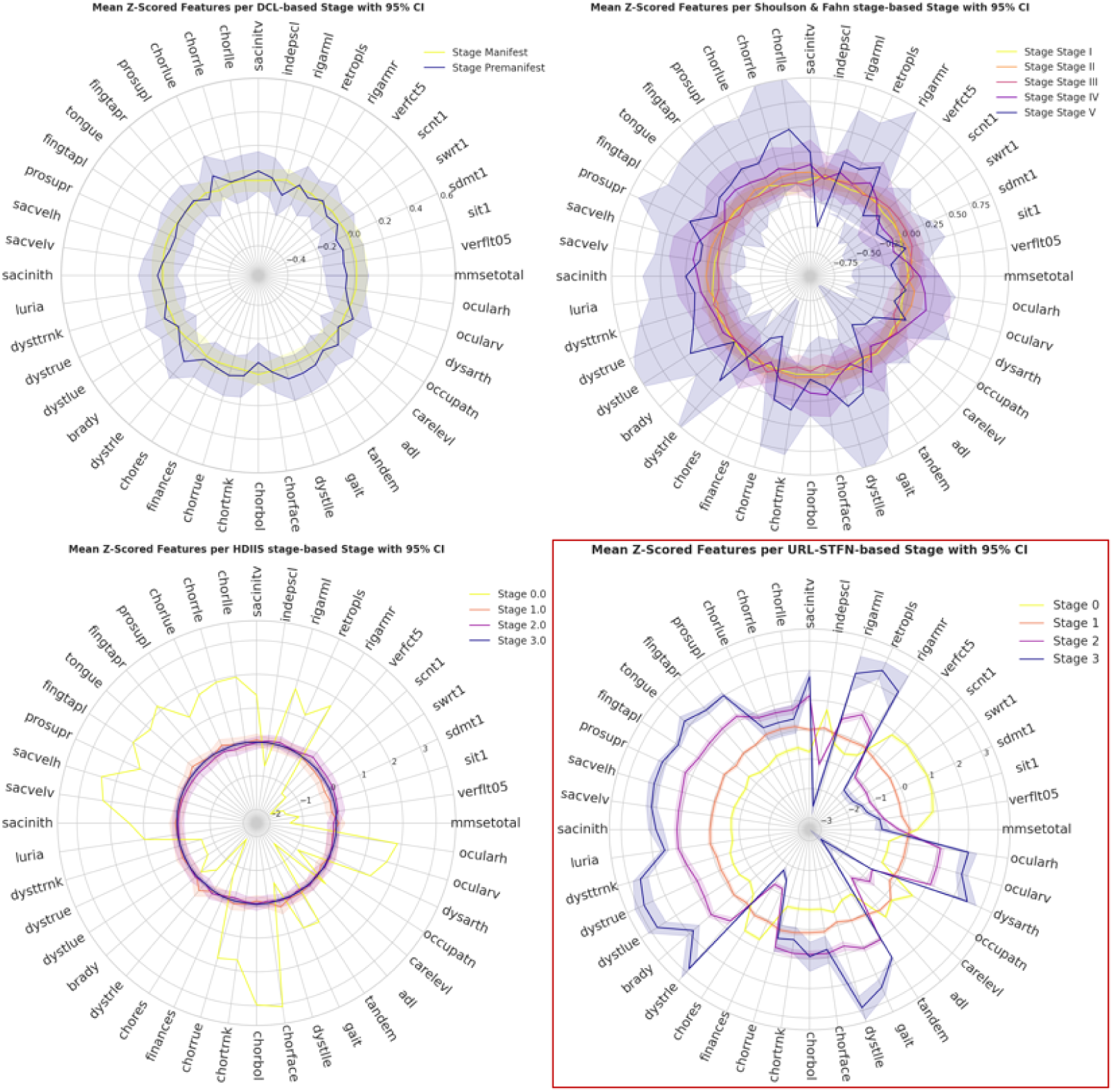
Radar plots for mean z-scored clinical feature profiles across four staging systems (colored lines): URL-STFN stages (outlined in red), HDISS stages (bottom left plot), TFC-based stages (top right plot), and DCL-based stages (top left plot). Each radar plot shows the mean of 44 standardized features for each stage, with shaded areas representing the 95% confidence intervals to illustrate feature dispersion. Feature labels are aligned along their respective axes for clarity.

**Figure 6:**
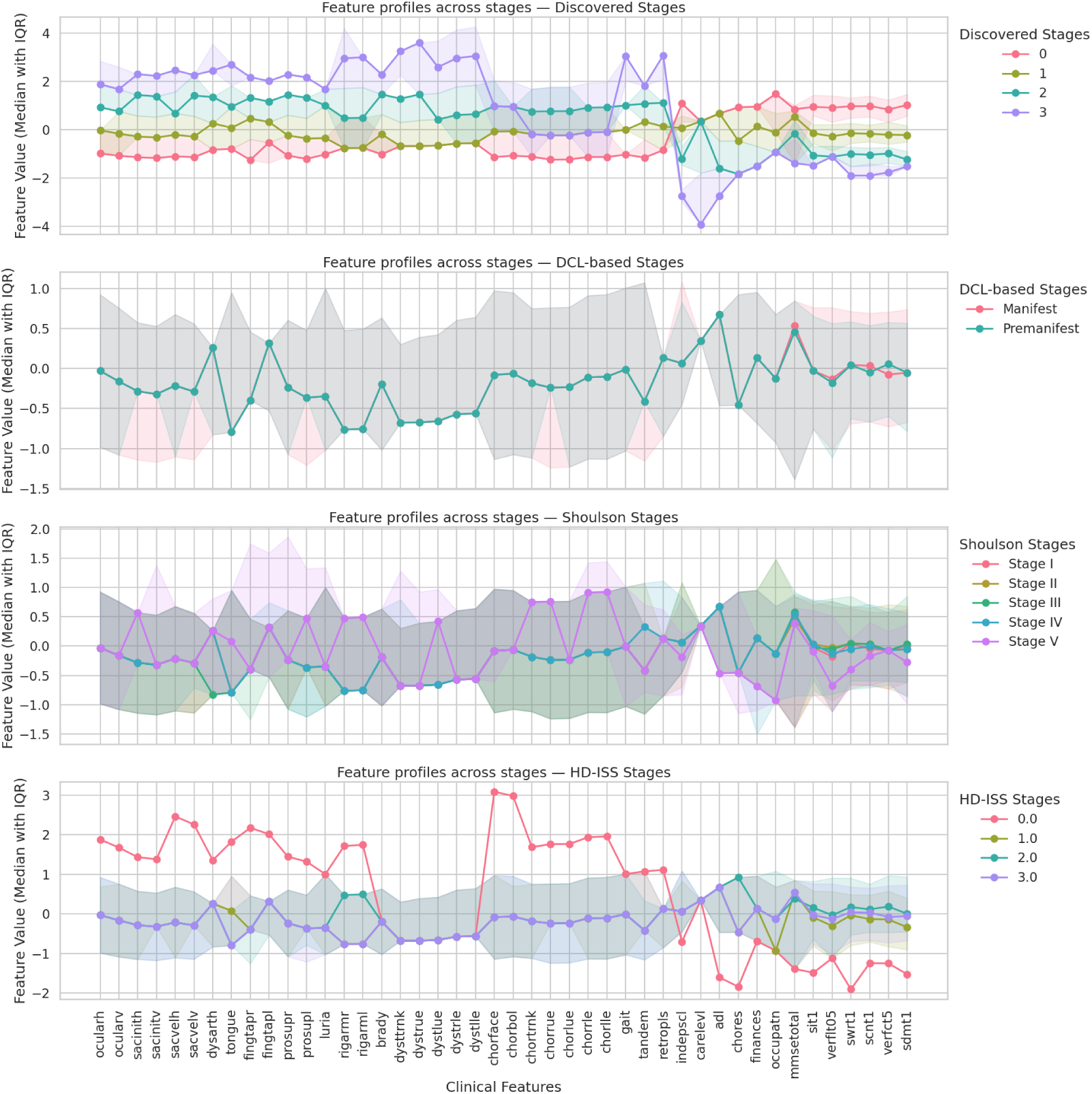
Line plots for the standardized median clinical feature values across the URL-STFN-based stages (Top graph), and conventional HD staging systems named in the figure as HDISS, TFC-based, and DCL-based stages (ordered from bottom to top plot, respectively).

The clearest separation of the discovered stages was also characterized by the monotonic shifts in median values and minimal interquartile overlap between adjacent clusters. This was particularly evident in cognitive measures, where SDMT1 mean and [IQR] declined from 44 [36–52] in Cluster 0 to 23 [17–29], 9 [5–15], and 0 [0-12.75] across Clusters 1–3, with limited overlap between early and late stages. A similar pattern was observed for Stroop Word Reading (swrt1) scores (89 [75–100] to 13.50 [0-30.75]). Functional measures showed even sharper discrimination, with Independence Scale (indepscl) scores decreasing from 100.00 [85–100] in Cluster 0 to 25 [20–30] in Cluster 3, and activity of daily living (adl) median scores dropping from 3.00 to 0.00, indicating a marked transition toward dependency with minimal overlap in advanced stages. Motor features further contributed to stage differentiation. For example, tongue protrusion scores increased from 0.00 [0.00–0.00] in Cluster 0 to 4.00 [3.00–4.00] in Cluster 3, while trunk dystonia and arm rigidity showed similar progressive patterns with limited overlap in later stages (Figure 6; Table D, Supplementary_File_2).

In contrast, conventional staging systems exhibited greater dispersion. DCL-based categories showed broader IQRs between premanifest and manifest stages, while HD-ISS and TFC-based staging demonstrated clearer separation mainly in advanced disease but less granularity during early to intermediate transitions, a consistent pattern across both median–IQR representations and standardized mean profiles in Figures 5 and 6, respectively.

### 3.4. Evaluation of Markov Model Derived Latent States

The implemented DHMM identified (*k* = 10) as the optimal number of latent states, achieving the highest log-likelihood. Although the log-likelihood variability across repeated runs was relatively high (standard deviation = 380), this reflected the most consistent performance among the tested values of *k*. The corresponding mean BIC was dropped to 237,532.92, which, despite being high, was the appropriate achieved balance between model fit and complexity relative to other candidate values of *k*. The agreement between the discovered latent states and established clinical stage proxies was comparable to the performance of the clustered URL-STFN latent representations. Specifically, the ARI was 0.0024 when compared with HD DCL-based categories and 0.1639 when compared with TFC-based stages. Although all clinical features differed significantly across the discovered states (p*<*0.05), and the feature profiles were more distinctly separated than those observed under conventional HD staging systems, visual inspection of DHMM line plot (Figure D, Supplementary_File_1) showed that the mean feature values for states 5 and 7, as well as states 1 and 4, were not clearly distinguishable, with measurements belonging to one state overlapping with other states. Nevertheless, the overall features progression pattern appeared clinically coherent, with states 1 and 4 reflecting intermediate disease severity, states 5 and 7 representing more advanced stages, and state 8 corresponding to the most severe clinical measurements.

The state-space representations demonstrated overall moderate-to-strong separation across disease stages, with variability across domains (Figure D). Cognitive and functional features exhibited the clearest stratification, characterized by consistent ordering of median values and limited interquartile overlap between states. Motor features such as bradykinesia and gait showed progressive differentiation with moderate overlap, whereas chorea-related measures displayed greater overlap and non-monotonic patterns, particularly in intermediate states (clearly apparent in Figures 5 and 6.

Overall, both approaches demonstrated comparable alignment with clinical proxies across cognitive, functional, and motor domains. However, the URL-STFN pipeline produced more clearly differentiated stage profiles, characterized by reduced overlap and more consistent progression patterns across features. In contrast, the DHMM approach incurred substantially higher computational cost due to the integration of longitudinal visit sequences, grid search for the optimal number of hidden states, and the high-dimensional feature space. This is consistent with known time-complexity limitations of Hidden Markov Models in longitudinal settings [69].

## 4. Discussion

The present study demonstrates that URL-based representations of Enroll-HD data can uncover clinically meaningful disease stages in HD, even without relying on pre-defined clinical stages or a single functional domain. Despite being tested on a limited subset of the dataset, the discovered clusters distinguished multidomain features—including cognitive, functional, and motor measures—more clearly than conventional staging approaches. Notably, the model revealed intermediate severity profiles within manifest disease, which are typically not distinguished in standard staging systems. This separation occurred without supervision from clinical experts or existing clinical stage labels, such as the DCL- or TFC-based staging, suggesting that the learned representations intrinsically encode severity patterns that are clinically interpretable.

The observed progression across clusters was consistent across multiple domains, as illustrated in Figure 4 and Table 3. Cognitive measures, including MMSE, SDMT, Stroop, and verbal fluency, demonstrated progressive deterioration from early to late clusters. In parallel, motor impairments, most notably bradykinesia, gait disturbances, postural instability, and oculomotor abnormalities, with functional deficits. Functional indices, particularly the Independence Scale and activities of daily living, declined sharply across clusters, reflecting the transition from partial independence to sustained dependency. This alignment of cognitive, motor, and functional decline underscores the multidomain nature of HD progression and highlights the increasing care requirements associated with advancing disease severity. Within this broader progression, specific motor features exhibited clinically meaningful patterns. Hyperkinetic chorea intensified in early to intermediate clusters but plateaued or attenuated in later stages, consistent with observations that chorea predominates in initial manifest stages before being replaced by bradykinesia, rigidity, and dystonia [70]. Similarly, oculomotor deficits, particularly impaired saccadic velocity and initiation, emerged early and progressively worsened, consistent with prior literature [71]. These domain-specific patterns not only validate the multidomain cluster separation but also reinforce that the discovered stages correspond to clinically recognizable disease phenomena.

The reduced overlap and consistent separation observed across the URL-STFN clusters provide strong support for their clinical validity. Unlike conventional staging systems, which rely on single-domain thresholds, the proposed framework integrates multidomain information, enabling finer stratification of disease severity, particularly in early and intermediate stages where clinical boundaries remain less well defined. The observed progression aligns with established knowledge of HD. Importantly, the model demonstrated clear separation even for motor features known to have limited inter-rater reliability, such as tongue protrusion and dystonia [62], suggesting robustness to clinical variability (see Figure 6). While both the URL-STFN and DHMM approaches demonstrated comparable alignment with clinical measures, the URL-STFN framework yielded more distinct and internally coherent stage profiles, with reduced overlap across disease severity levels. In addition, it required substantially less computational time, making it more practical for large-scale datasets such as Enroll-HD.

These findings suggest that representation-based unsupervised learning provides a more efficient and scalable framework for disease stage discovery compared to traditional sequence-based models. Nevertheless, DHMMs remain valuable for capturing temporal dynamics, and future work should explore HMM-based or hybrid approaches to further refine stage characterization.

The meaningful patterns extracted by the proposed URL-STFN-based representation approach outperform conventional deep learning latent feature discovery methods, and when clustered through the unsupervised learning pipeline, enhance current classification systems by providing a more granular and objective characterization of HD stages. This framework enables out-of-sample stage assignment, by allowing individual patients to be mapped onto the existing disease stage structure without retraining the model by processing patient’s clinical features using the same preprocessing and feature construction pipeline as the training data, and then projecting them into the learned representation space of the URL-STFN model. The resulting latent representation can subsequently be assigned to a disease stage based on proximity to the learned cluster centroids (or corresponding stage prototypes). Consequently, the framework could improve patient stratification in clinical trials, support individualized prognosis, and inform clinical care planning. Collectively, this work offers proof-of-principle validation that unsupervised representation learning can identify clinically meaningful disease stages in HD, producing results consistent with established clinical knowledge while also detecting subtler, intermediate phenotypes that conventional methods may overlook.

## 5. Limitation and Future Work

Several limitations of the present study should be acknowledged. First, the proposed framework adopts a transductive learning setting in which all patients are incorporated during graph construction and model training. Although the evaluation protocol employed bootstrapping procedures to reduce overfitting and improve robustness, the framework has not yet been evaluated in a fully inductive setting where the stages are generated for entirely unseen patients. Such validation could provide stronger evidence regarding the model’s generalizability and clinical applicability. Future work will therefore focus on evaluating the framework on independent unseen datasets and exploring inductive graph learning approaches suitable for deployment in real-world clinical settings.

A second limitation concerns the use of absolute age as the primary dynamic graph anchor, assuming that disease progression patterns are sufficiently aligned by chronological age alone. Another option would be to use relative temporal measures, such as time since disease onset or diagnosis. However, deriving reliable relative temporal anchors remains challenging due to heterogeneity of the disease progression inconsistencies and incomplete longitudinal clinical records. Areas to explore in future work include varying age-bucket granularities, as well as exploring novel extensions of the graph convolution framework to operate on a three-dimensional adjacency representation capable of jointly modelling temporal, structural, and feature-level relationships.

The study was also conducted on a relatively small subset of the Enroll-HD dataset with limited longitudinal coverage. Consequently, the discovered progression patterns and patient clusters may not fully capture the heterogeneity of the broader HD population. Future studies will extend the analysis to larger cohorts with longer longitudinal follow-up periods and more comprehensive clinical records to improve external validity and robustness. We will also incorporate morphometric and biofluid biomarkers which could further enhance the objectivity and robustness of the discovered HD staging framework. Finally, model interpretability remains an important challenge in graph-based deep learning applications for healthcare. While the proposed framework demonstrated promising clustering and representation capabilities, the specific clinical features contributing to the learned patient representations, cluster formation, and progression transitions remain incompletely understood. Future work will focus on investigating explainable graph learning approaches to better identify the clinical variables and graph relationships driving cluster assignments and temporal transition patterns.

## Data Availability

All data analyzed in this study were obtained from the CHDI Foundation through the Enroll-HD platform. The data are not publicly available and are subject to the CHDI Foundation's data access policies. Researchers interested in obtaining the same analyzed dataset should contact the corresponding author for guidance on the data access process.

https://enroll-hd.org/

## Acknowledgements

Data used in this work was generously provided by the participants in the Enroll-HD study and made available by CHDI Foundation, Inc. Enroll-HD is a global clinical research platform intended to accelerate progress towards therapeutics for Huntington’s disease; core datasets are collected annually on all research participants as part of this multi-center longitudinal observational study. Enroll-HD is sponsored by CHDI Foundation, Inc., a nonprofit biomedical research organization exclusively dedicated to developing therapeutics for Huntington’s disease. Enroll-HD would not be possible without the vital contribution of the research participants and their families. [61].

## Data Availability and Supplementary Materials

The Enroll-HD dataset used in the assessment of the proposed graph model can be requested directly from Enroll-HD clinical research platform; https://www.enroll-hd.org/. For code access and work reproducibility, contact the corresponding author. The Excel sheet of the supplementary file contains the list of features used, the name of the source table in the Enroll-HD main dictionary file, the types of variables and the descriptions.

## Ethics statement

This study used secondary, de-identified data from the Enroll-HD cohort provided under the governance of the CHDI Foundation. All data access complied with the Enroll-HD study procedures and relevant ethical approvals obtained by the original study. No additional data was collected, and no direct participant participation occurred in this work. Therefore, no new ethical approval or informed consent was required for this analysis.

## Author contributions

Authors’ initials and roles according to the CRediT taxonomy are:

L.M.A.Z.: Conceptualization; Methodology; Investigation; Data Curation; Formal Analysis; Validation; Writing - Original Draft.

H.Z.: Writing – Review&Editing.

A.G.: Writing – Review&Editing.

J.R.W.: Writing – Review&Editing.

M.L.: Validation; Writing – Review&Editing.

M.V.: Writing – Review&Editing.

