## Supplementary material for "Dynamic Graph Representation Learning for Data-Driven Huntington’s Disease Staging: Evaluation Against Existing Embedding Methods and State-Space Models": Supplementary_File_1.docx

**Table A.** Summary of Enroll-HD Variables: This table lists key, motor, functional, cognitive, and post-hoc variables (the underlined) from the enroll table , including the variable name, label, category, data type, source file, and full coding description. All features were used in the representation model to uncover disease stages, except those indicated for post-hoc analysis.

| **Variable** | **Label** | **Category** | **Type** | **Description (full coding)** |
| --- | --- | --- | --- | --- |
| age | Age at visit | Key | Numeric | Age at visit (years). |
| capscore | CAP score | Key | Numeric | CAP score = Age × (CAG − 30) / 6.49. |
| motscore | Total Motor Score (TMS) | Motor | Numeric | Sum of all UHDRS motor items; computed when all items are scored. |
| ocular | Ocular pursuit – Horizontal | Motor | Ordinal | 0=complete (normal); 1=jerky movement; 2=interrupted pursuits/full range; 3=incomplete range; 4=cannot pursue. |
| ocular | Ocular pursuit – Vertical | Motor | Ordinal | 0=complete (normal); 1=jerky movement; 2=interrupted pursuits/full range; 3=incomplete range; 4=cannot pursue. |
| sacinith | Saccade initiation – Horizontal | Motor | Ordinal | 0=normal; 1=increased latency only; 2=suppressible blinks/head movements; 3=unsuppressible head movements; 4=cannot initiate saccades. |
| sacinitv | Saccade initiation – Vertical | Motor | Ordinal | 0=normal; 1=increased latency only; 2=suppressible blinks/head movements; 3=unsuppressible head movements; 4=cannot initiate saccades. |
| sacvelh | Saccade velocity – Horizontal | Motor | Ordinal | 0=normal; 1=mild slowing; 2=moderate slowing; 3=severely slow (full range); 4=incomplete range. |
| sacvelv | Saccade velocity – Vertical | Motor | Ordinal | 0=normal; 1=mild slowing; 2=moderate slowing; 3=severely slow (full range); 4=incomplete range. |
| dysarth | Dysarthria | Motor | Ordinal | 0=normal; 1=unclear (no repetition needed); 2=must repeat to be understood; 3=mostly incomprehensible; 4=anarthria. |
| tongue | Tongue protrusion | Motor | Ordinal | 0=can hold fully protruded for 10 sec; 1=cannot hold for 10 sec; 2=cannot hold for 5 sec; 3=cannot fully protrude; 4=cannot protrude beyond lips. |
| fingtapr | Finger taps – Right | Motor | Ordinal | 0=normal (≥15/5 sec); 1=mild slowing (11–14); 2=moderate (7–10); 3=severe (3–6); 4=barely performs (0–2). |
| fingtapl | Finger taps – Left | Motor | Ordinal | Same coding as right hand. |
| prosupr | Pronate/supinate – Right | Motor | Ordinal | 0=normal; 1=mild slowing/irregular; 2=moderate; 3=severe; 4=cannot perform. |
| prosupl | Pronate/supinate – Left | Motor | Ordinal | Same coding as right hand. |
| luria | Luria sequence | Motor | Ordinal | 0≥4/10 sec no cue; 1<4 no cue; 2≥4 with cue; 3<4 with cue; 4=cannot perform. |
| rigarmr | Rigidity – Right arm | Motor | Ordinal | 0=absent; 1=slight/with activation; 2=mild–moderate; 3=severe full range; 4=severe limited range. |
| rigarml | Rigidity – Left arm | Motor | Ordinal | Same coding as right arm. |
| brady | Bradykinesia | Motor | Ordinal | 0=normal; 1=minimally slow; 2=mild; 3=moderate; 4=markedly slow with delayed initiation. |
| gait | Gait | Motor | Ordinal | 0=normal; 1=wide/slow; 2=wide with difficulty; 3=walks with assistance; 4=cannot attempt. |
| tandem | Tandem walking | Motor | Ordinal | 0=normal; 1=1–3 deviations; 2>3 deviations; 3=cannot complete; 4=cannot attempt. |
| retropls | Retropulsion test | Motor | Ordinal | 0=normal; 1=recovers spontaneously; 2=would fall if not caught; 3=tends to fall; 4=cannot stand. |
| diagconf | Diagnostic Confidence Level (DCL) | Motor | Ordinal | 0=normal; 1<50% confidence; 2=50–89%; 3=90–98%; 4≥99% (manifest HD). |
| tfcscore | Total Functional Capacity (TFC) | Functional | Numeric | Range 0–13; higher indicates better functional capacity. |
| occupatn | Occupation | Functional | Ordinal | 0=unable; 1=marginal work; 2=reduced capacity; 3=normal. |
| finances | Finances | Functional | Ordinal | 0=unable; 1=major assistance; 2=slight assistance; 3=normal. |
| chores | Domestic chores | Functional | Ordinal | 0=unable; 1=impaired; 2=normal. |
| adl | Activities of Daily Living | Functional | Ordinal | 0=total care; 1=gross tasks only; 2=minimal impairment; 3=normal. |
| carelevl | Care level | Functional | Ordinal | 0=skilled nursing; 1=home/chronic care; 2=home. |
| indepscl | Independence scale (%) | Functional | Ordinal | A multiple of 5; 0–100% in increments (e.g., 100=no care needed; 90–70 varying independence; ≤50 increasing assistance; 0–10 total bed care). |
| sdmt1 | SDMT total correct | Cognitive | Numeric | Total correct responses (processing speed test). |
| verfct5 | Verbal fluency (category) | Cognitive | Numeric | Total correct words generated in 1 minute. |
| scnt1 | Stroop color naming | Cognitive | Numeric | Total correct responses. |
| swrt1 | Stroop word reading | Cognitive | Numeric | Total correct responses. |
| sit1 | Stroop interference | Cognitive | Numeric | Total correct responses under interference condition. |
| verflt05 | Letter verbal fluency | Cognitive | Numeric | Total correct words generated in 3 minutes. |
| mmsetotal | MMSE total score | Cognitive | Numeric | Score range 0–30; higher indicates better cognitive function. |
| dysttrnk | Dystonia trunk | Motor | Ordinal | absent:0; slight/intermittent:1; mild/common or moderate/intermittent:2; moderate/common:3; marked/prolonged:4 |
| dystrue | Dystonia right upper extremity | Motor | Ordinal | absent:0; slight/intermittent:1; mild/common or moderate/intermittent:2; moderate/common:3; marked/prolonged:4 |
| dystlue | Dystonia left upper extremity | Motor | Ordinal | absent:0; slight/intermittent:1; mild/common or moderate/intermittent:2; moderate/common:3; marked/prolonged:4 |
| dystrle | Dystonia right lower extremity | Motor | Ordinal | absent:0; slight/intermittent:1; mild/common or moderate/intermittent:2; moderate/common:3; marked/prolonged:4 |
| dystlle | Dystonia left lower extremity | Motor | Ordinal | absent:0; slight/intermittent:1; mild/common or moderate/intermittent:2; moderate/common:3; marked/prolonged:4 |
| chorface | Chorea face | Motor | Ordinal | absent:0; slight/intermittent:1; mild/common or moderate/intermittent:2; moderate/common:3; marked/prolonged:4 |
| chorbol | Chorea body | Motor | Ordinal | absent:0; slight/intermittent:1; mild/common or moderate/intermittent:2; moderate/common:3; marked/prolonged:4 |
| chortrnk | Chorea trunk | Motor | Ordinal | absent:0; slight/intermittent:1; mild/common or moderate/intermittent:2; moderate/common:3; marked/prolonged:4 |
| chorrue | Chorea right upper extremity | Motor | Ordinal | absent:0; slight/intermittent:1; mild/common or moderate/intermittent:2; moderate/common:3; marked/prolonged:4 |
| chorlue | Chorea left upper extremity | Motor | Ordinal | absent:0; slight/intermittent:1; mild/common or moderate/intermittent:2; moderate/common:3; marked/prolonged:4 |
| chorrle | Chorea right lower extremity | Motor | Ordinal | absent:0; slight/intermittent:1; mild/common or moderate/intermittent:2; moderate/common:3; marked/prolonged:4 |
| chorlle | Chorea left lower extremity | Motor | Ordinal | absent:0; slight/intermittent:1; mild/common or moderate/intermittent:2; moderate/common:3; marked/prolonged:4 |
| ***Notes.***  *Variables or clinical features parameters and the coding of these parameters can be found in enroll-hd data dictionary.  https://enroll-hd.org/for-researchers/data-support-documentation/#documentation/doc-details2/68b60a0a59371b02d0740572/kn-asset/103-206-137-68b609da96de8802d6a30de6/enrollhd_datadictionary_20250807.xlsx | | | | |

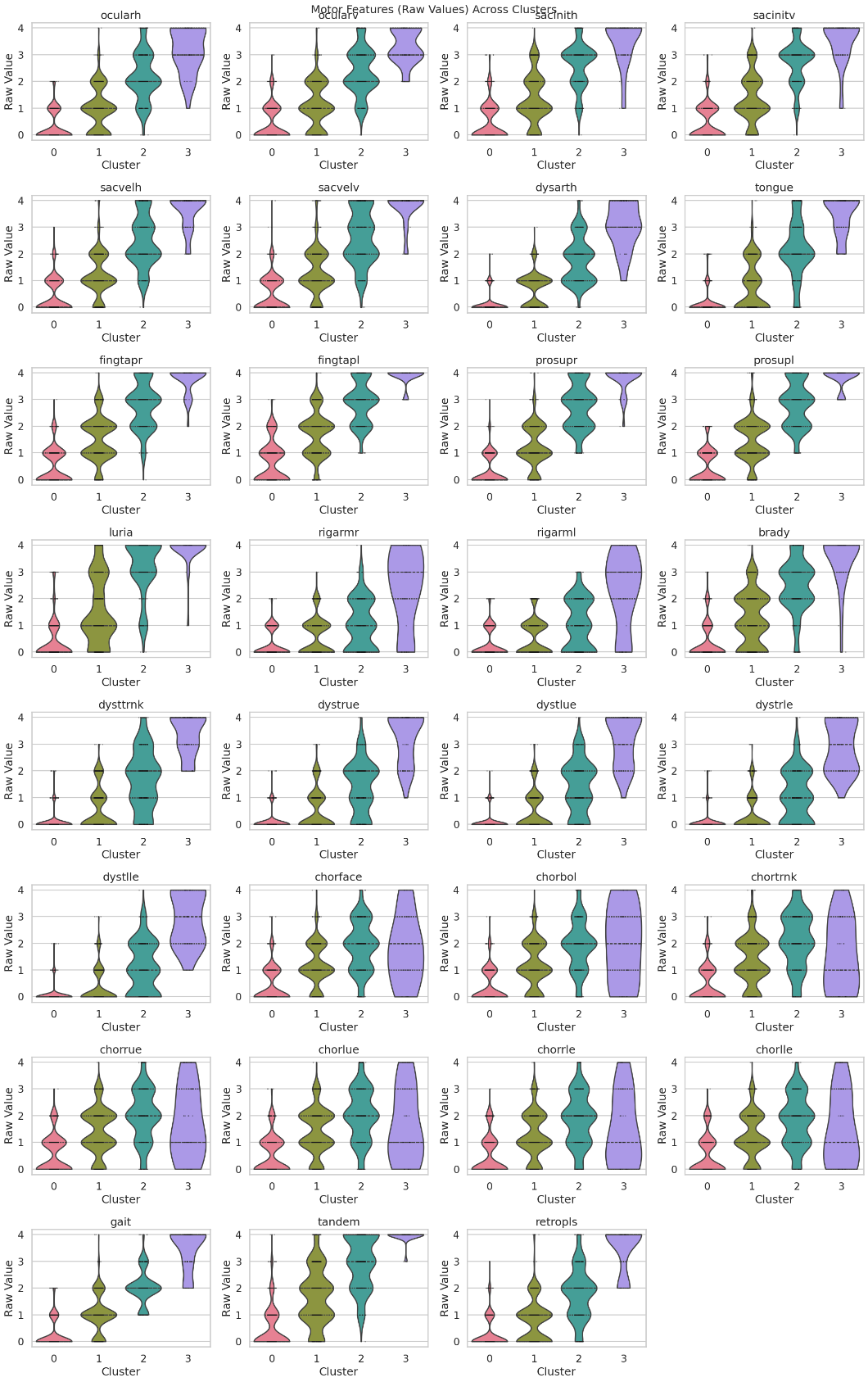

**Figure A.** Violin plots of motor features across the URL-STFN based discovered stages (Clusters 0–3) in Huntington's disease. All the measures displayed were statistically significant across stages, and the distributions demonstrate progressive motor incline with advancing discovered stage.

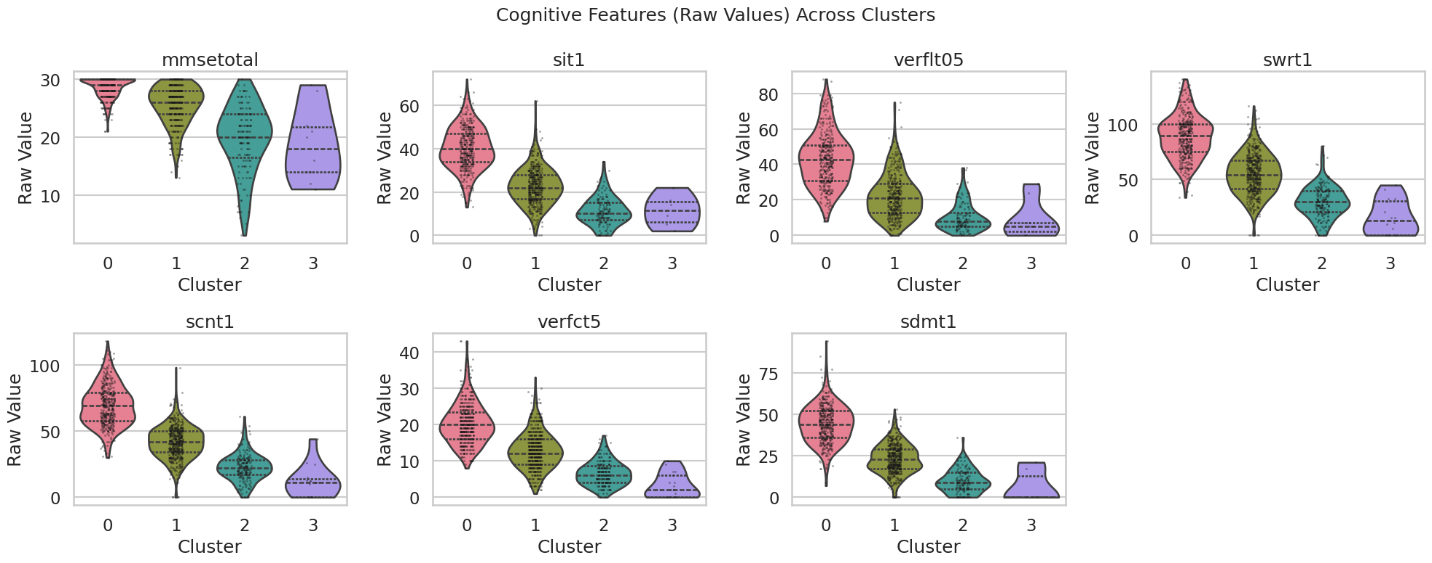

**Figure B.** Violin plots of cognitive features across the URL-STFN based discovered stages (Clusters 0–3) in Huntington's disease. Displayed measures were statistically significant across stages, and their distributions demonstrate progressive functional decline with advancing discovered stage.

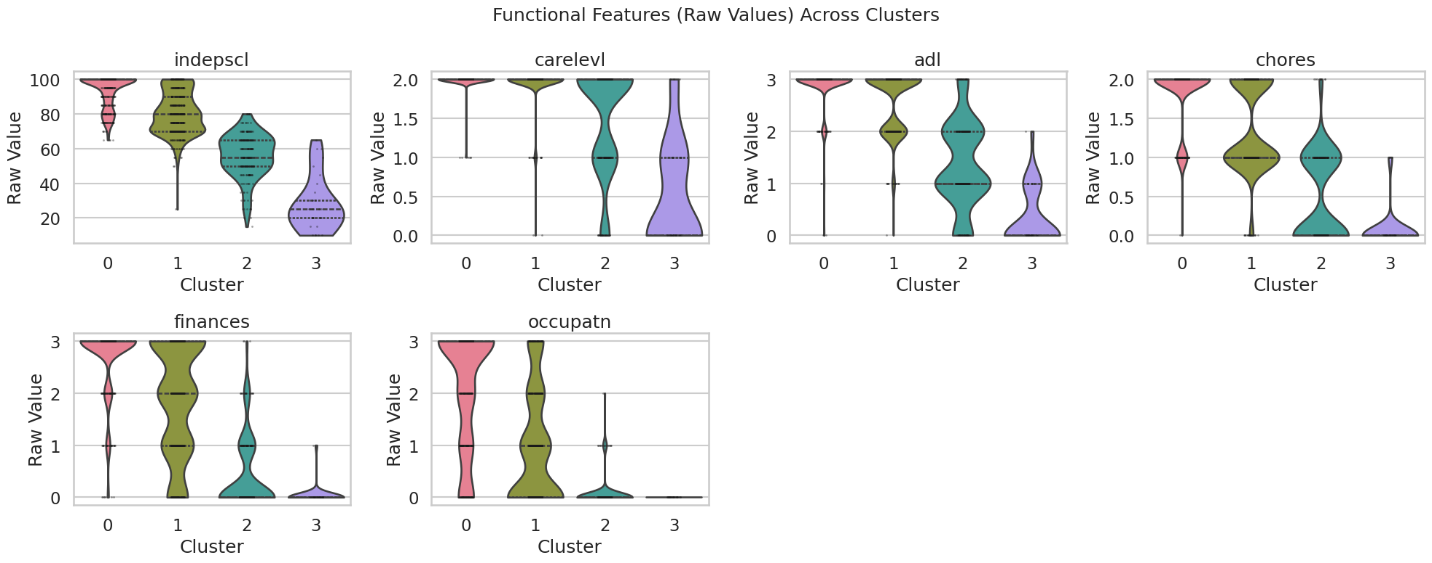

**Figure C.** Violin plots of functional features across the URL-STFN based discovered stages (Clusters 0–3) in Huntington's disease. Displayed measures were statistically significant across stages, and their distributions demonstrate progressive cognitive decline with advancing discovered stage.

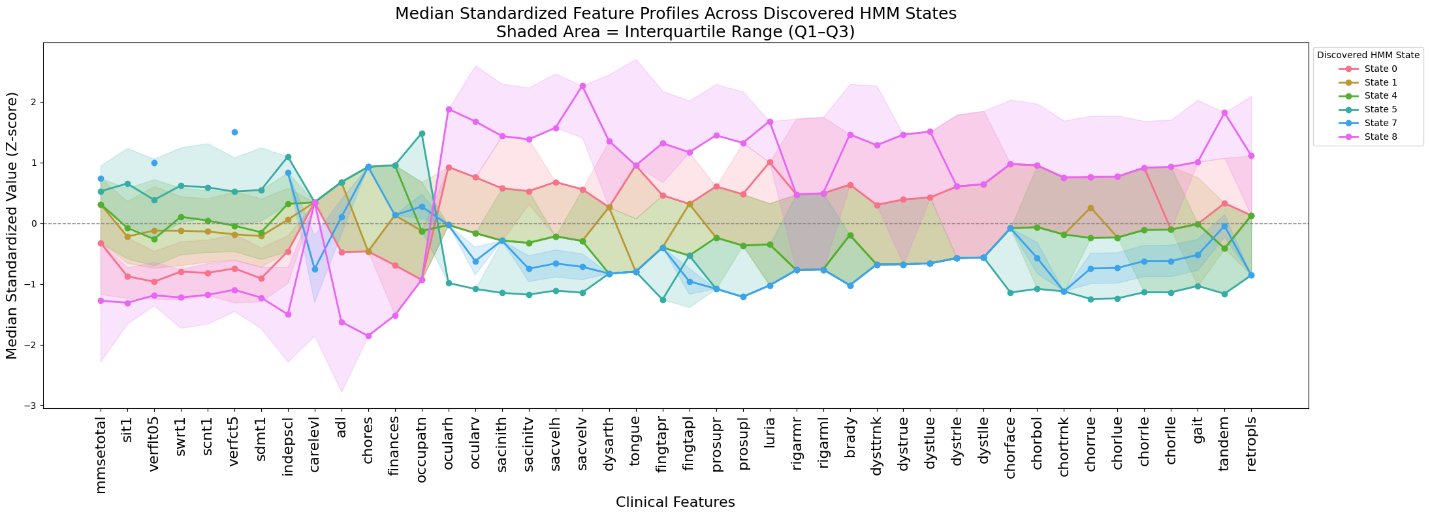

**Figure D.** The median of standardized values of clinical features across different HMM discovered states. Each line represents a distinct HMM state. Note that the HMM model was trained with 10 latent states, each with distinct emission probabilities. During state decoding (using model.predict()), each observation is assigned the single most likely state, and some states may be very specialized or rare. Consequently, not every state is necessarily assigned to the clinical visit points, which explains why only 6 of the 10 states appear in the merged dataset and this plot.

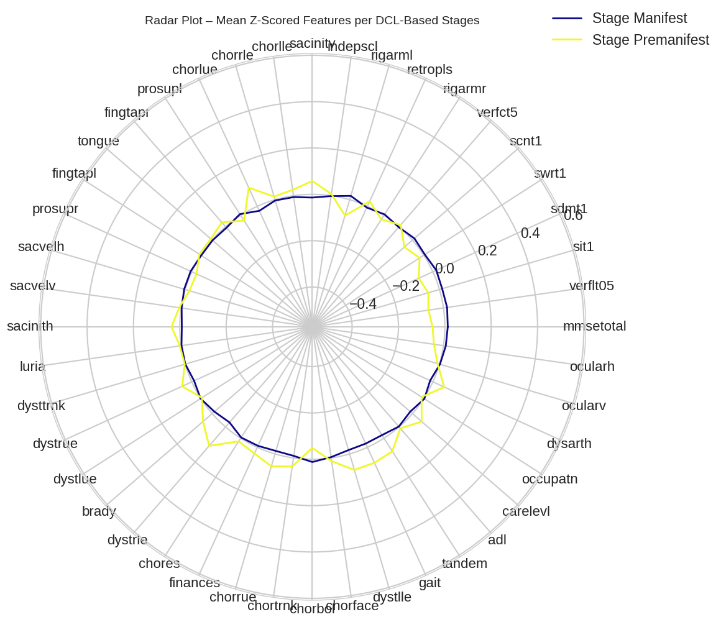

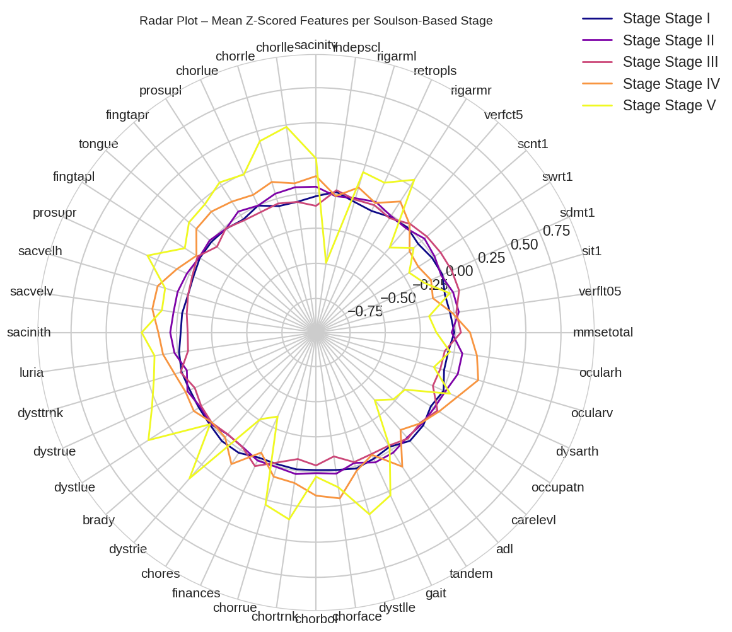

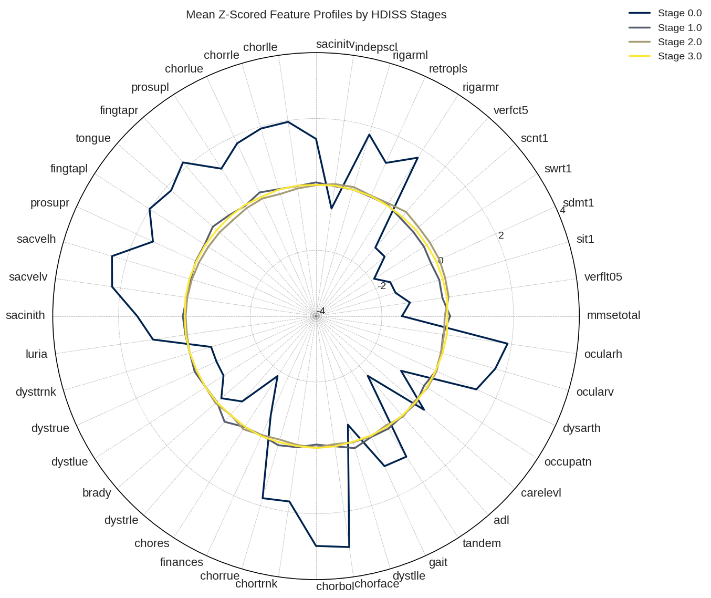

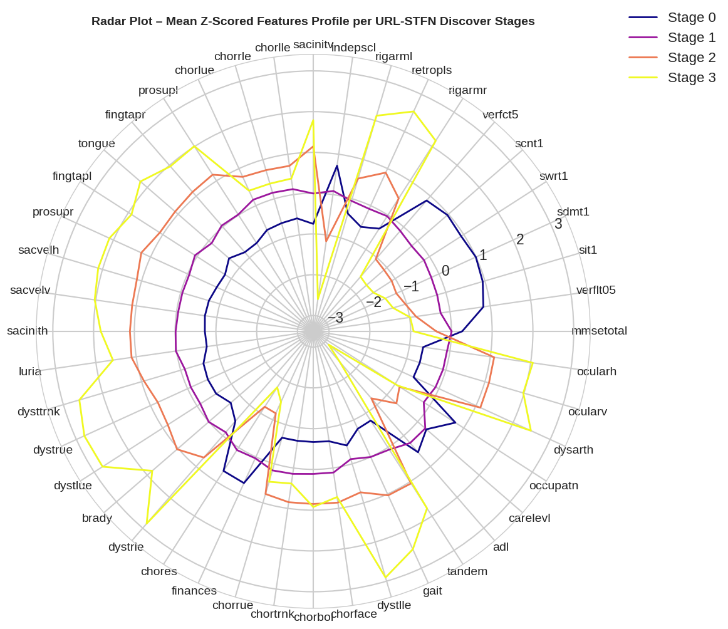

**Figure E.** Comparison of motor and cognitive feature profiles across different Huntington’s disease staging systems. The radar plots show the mean z-scored values of 44 clinical features for each stage. Top left: DCL-based stages; bottom left: HDISS stages; top right: Shoulson-based stages. Bottom right: URL-STFN discovered clusters, with cluster boundaries outlined in red. Each polygon represents the mean profile of a stage or cluster, allowing visual comparison of feature distributions across staging systems.
